# Corporate activities that influence population health: A scoping review and qualitative synthesis to develop the HEALTH-CORP typology

**DOI:** 10.1101/2024.04.09.24305564

**Authors:** Raquel Burgess, Kate Nyhan, Nicholas Freudenberg, Yusuf Ransome

## Abstract

**Introduction:** The concept of the commercial determinants of health (CDH) is used to study the actions (and associated structures) of commercial entities that influence population health and health equity. The aim of this study was to develop a typology that describes the diverse set of activities through which corporations influence population health and health equity across industries.

**Methods:** We conducted a scoping review of articles using CDH terms (n=116) that discuss corporate activities that can influence population health and health equity across 16 industries. We used the qualitative constant comparison method to build a typology called the Corporate Influences on Population Health (HEALTH-CORP) typology.

**Results:** The HEALTH-CORP typology identifies 70 corporate activities that can influence health across industries and categorizes them into seven domains of corporate influence (e.g., political practices, employment practices). We present a model that situates these domains based on their proximity to health outcomes and identify five population groups (e.g., workers, local communities) to consider when evaluating corporate health impacts.

**Discussion:** The HEALTH-CORP typology facilitates an understanding of the diverse set of corporate activities that can influence population health and the population groups affected by these activities. We discuss the utility of these contributions in terms of identifying interventions to address the CDH and advancing efforts to measure and monitor the CDH. We also leverage our findings to identify key gaps in CDH literature and suggest avenues for future research.

## Introduction

For centuries, commercial actors and the structures that govern them have shaped the health of various populations in profound ways [1]. Growing attention to this influence recognizes the increasing economic, political, and legal power wielded by corporations, especially those that operate transnationally. In the past two decades, scholars have studied this issue through the lens of the ‘commercial determinants of health’ (CDH). In a recent *Lancet*-commissioned series on the topic, the CDH were defined as “the systems, practices, and pathways through which commercial actors drive health and equity” [2].

Gilmore and colleagues [2] provide us with a detailed summary of the CDH literature to-date and proposed a conceptual model for understanding how the CDH influence population health outcomes. Drawing on existing models of the political and social determinants of health (SDH), their model highlights the ways that corporate practices (e.g., political practices, financial practices) exert influence on the political and economic system, which have downstream impacts on SDH such as housing, labour, as well as on the natural environment. Gilmore and colleagues highlight how the maldistribution of power between the private and public sector creates current “pathological” economic and political systems that favour the interests of commercial entities, often at the expense of health and equity [2].

We conducted a scoping review and qualitative synthesis of 116 CDH articles discussing corporate activities with the potential to influence population health and health equity. Leveraging the findings from this process, we offer three distinct contributions that add to existing literature [2–8]. First, we develop a typology (called the Corporate Influences on Health (HEALTH-CORP) typology) that identifies 70 specific corporate activities with the potential to influence human health across industries and categorizes these activities into seven domains of corporate influence (e.g., political practices, products and services). Second, we situate the domains based on their proximity to health outcomes (i.e., distal to proximal) and propose five population groups to consider when evaluating the health impact of corporate practices. Our third contribution is to use our findings to illuminate current gaps in CDH research.

In the following sections, we report our review methods and describe how we used the literature to build the HEALTH-CORP typology. We summarize the activities we identified as relevant to the seven domains of influence and five population groups. We conclude by discussing the utility of these contributions and identifying avenues for future CDH research.

## Methods

We used scoping review methodology to find relevant literature and qualitative synthesis to build the typology. Scoping review methodology is preferred when examining emerging fields of research and consolidating key factors related to a particular concept [9]. Likewise, qualitative synthesis is useful for systematically interpreting research to identify and represent its meaning [10]. We report our methods and findings in accordance with the PRISMA extension for scoping reviews (PRISMA-ScR) [11]. No study protocol was registered.

### Search strategy

We consulted prior work by de Lacy-Vawdon and Livingstone [12] to build our search. We expanded the scope of prior searches for CDH terms ((commercial OR corporate) AND determinant* AND (health OR disease*)), to include related terms such as “corporate political activity” and “morbidity” (Additional File 1, Appendix 1). At the full-text screening stage, we then excluded articles that did not specifically mention CDH terms (i.e., “commercial determinants”, “corporate determinant(s)”). The purpose was to identify as many related articles as possible, including those that did not use CDH terms in their title, abstract, or keywords, but nevertheless used CDH terms in the full text. We searched Scopus, OVID Medline, Ovid Embase, and Ovid Global Health with no date restrictions on Jan 4, 2022 and again on Sept 13, 2022.

### Eligibility criteria

Eligible articles described activities (i.e., decisions, strategies, or other actions) that corporations or those acting on behalf of corporations engage in that have been demonstrated to or have the potential to influence population health and/or health equity. This criterion was applied broadly to identify as many relevant activities as possible; the potential to influence population health and/or health equity could have been investigated explicitly within the respective study, supported by previous research (for e.g., changes to income are known to influence health), or reasonably be expected to influence population health (e.g., delayed implementation of evidence-based health policy due to corporate influence). Moreover, the expected impact on health could be direct (e.g., occupational injury), indirect (e.g., decreased health protections due to corporate lobbying), health-promoting (e.g., provision of income), or health-adverse (e.g., harmful product), or some combination of these categories.

Articles were excluded if the full text was not available in English and if the title, abstract, keywords, or full text did not contain CDH terms (excluding the reference list). Full-length books were also excluded.

### Screening

Following manual and software-assisted (via Mendeley [13] reference management software) de-duplication, the first author (R.B.) screened the titles and abstracts in Rayyan [14], a web service for organizing reviews. We did not employ double, independent screening for verification purposes. Full texts were retrieved for eligible records. Then, EndNote’s [15] ‘Smart Group’ feature was used to screen for articles that use CDH terms.

### Data analysis

Using the qualitative data analysis software NVivo 12 [16], the first author (R.B) employed the constant comparative method to inductively code the data with the goal of identifying relevant corporate activities and arranging them into overarching descriptive domains [17,18].

Specifically, examples of corporate activities discussed in the included texts (e.g., corporate involvement in nutrition education in schools [19]) were coded and compared to other relevant examples (e.g., corporate provision of resources on breastfeeding for mothers [20]), to identify overarching activities (e.g., corporate involvement in health education directed at the public). These activities were then grouped into larger descriptive categories (e.g., preference and perception shaping practices), which we call ‘domains of corporate influence’. In the final typology, specific activities were included if they had been described in a general sense in the included literature or discussed in relation to two or more industries.

Drawing on the findings of this analysis, other CDH work [2,3,6,7], and the work of relevant civil society organizations [21,22], the first author (R.B.) then developed a model to describe the interdependencies between domains and their proximity to health outcomes in consultation with the third author (N.F.). This included identifying five population groups whose health is affected by corporate activities (i.e., consumers, workers, disadvantaged groups, vulnerable groups, and local communities).

### Critical appraisal

Consistent with scoping review methodology [23], a formal quality assessment of articles was not performed.

## Results

### Article Screening

7910 records were identified during the initial search, 4924 of which remained after de-duplication. Following title and abstract screening, 430 full texts were sought and 412 were found. 248 articles were excluded based on screening for use of CDH terms. Of the remaining 164 articles, 74 were included. Another 42 articles were included when the search was conducted a second time for a total of 116 included articles (Figure 1).

**Figure 1.**
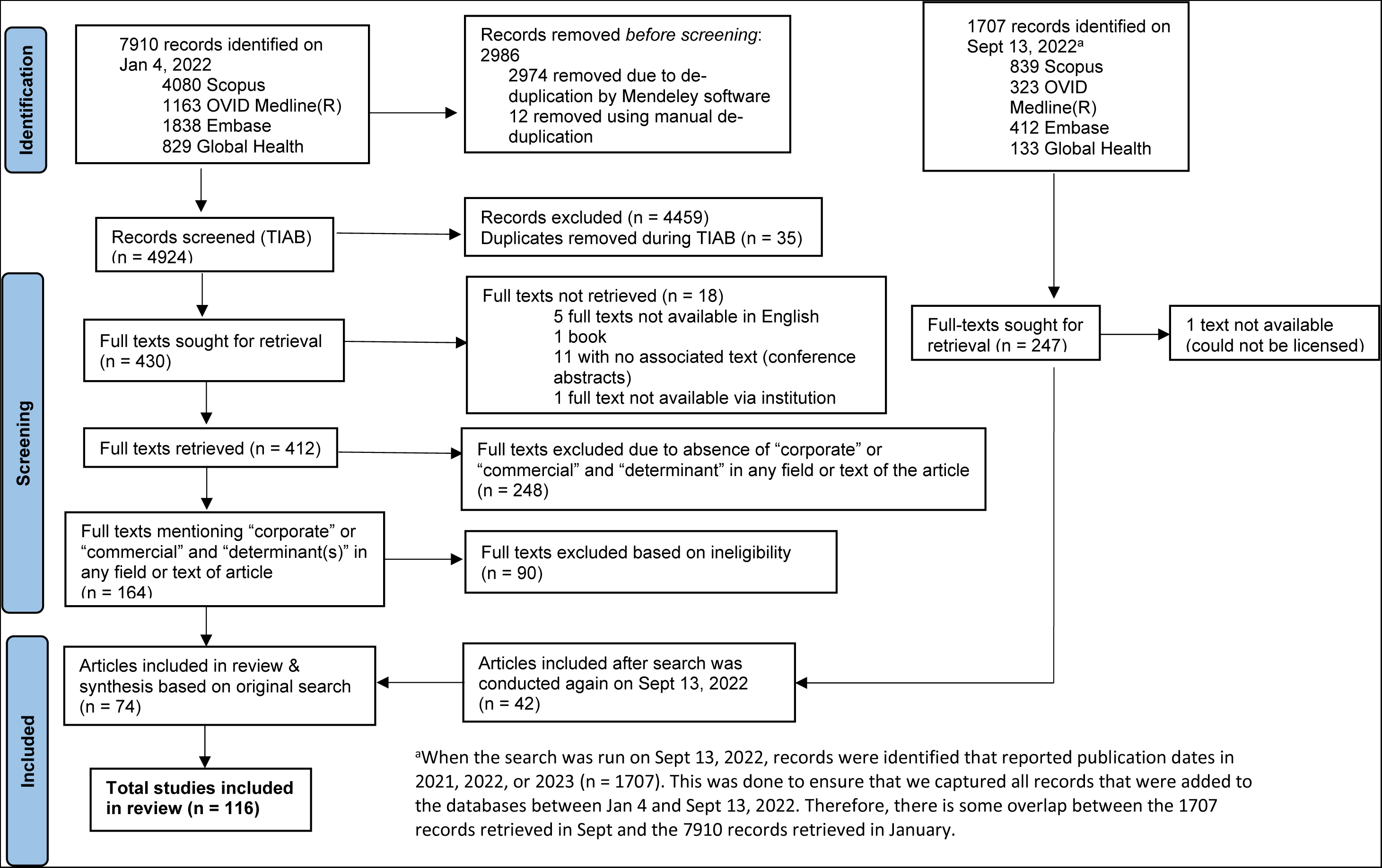
PRISMA Diagram.

### Characteristics of Included Articles

Almost half of the articles included were conceptual (50 articles; 43%) and a substantial proportion employed qualitative methods (37; 32%). Fifty eight percent (67 articles) focused on the food and beverage, tobacco, and alcohol industries, with less research dedicated to studying other industries (e.g., social media, gambling, fossil fuels). Of the 58 included articles that reported a regional focus, 72% (42 articles) studied corporate activities in high-income countries (HICs), such as United States, United Kingdom, or Australia. Detailed descriptions of the characteristics of the included literature are reported in our previous work [24]. Individual article characteristics are provided in Additional File 1, Appendix 2.

### Corporate Influences on Population Health (HEALTH-CORP)

We identified seven domains of corporate influence, which are: 1) political practices, 2) preference and perception shaping practices, 3) corporate social responsibility practices, 4) economic practices, 5) products & services, 6) employment practices, and 7) environmental practices (Table 1).

**Table 1.**
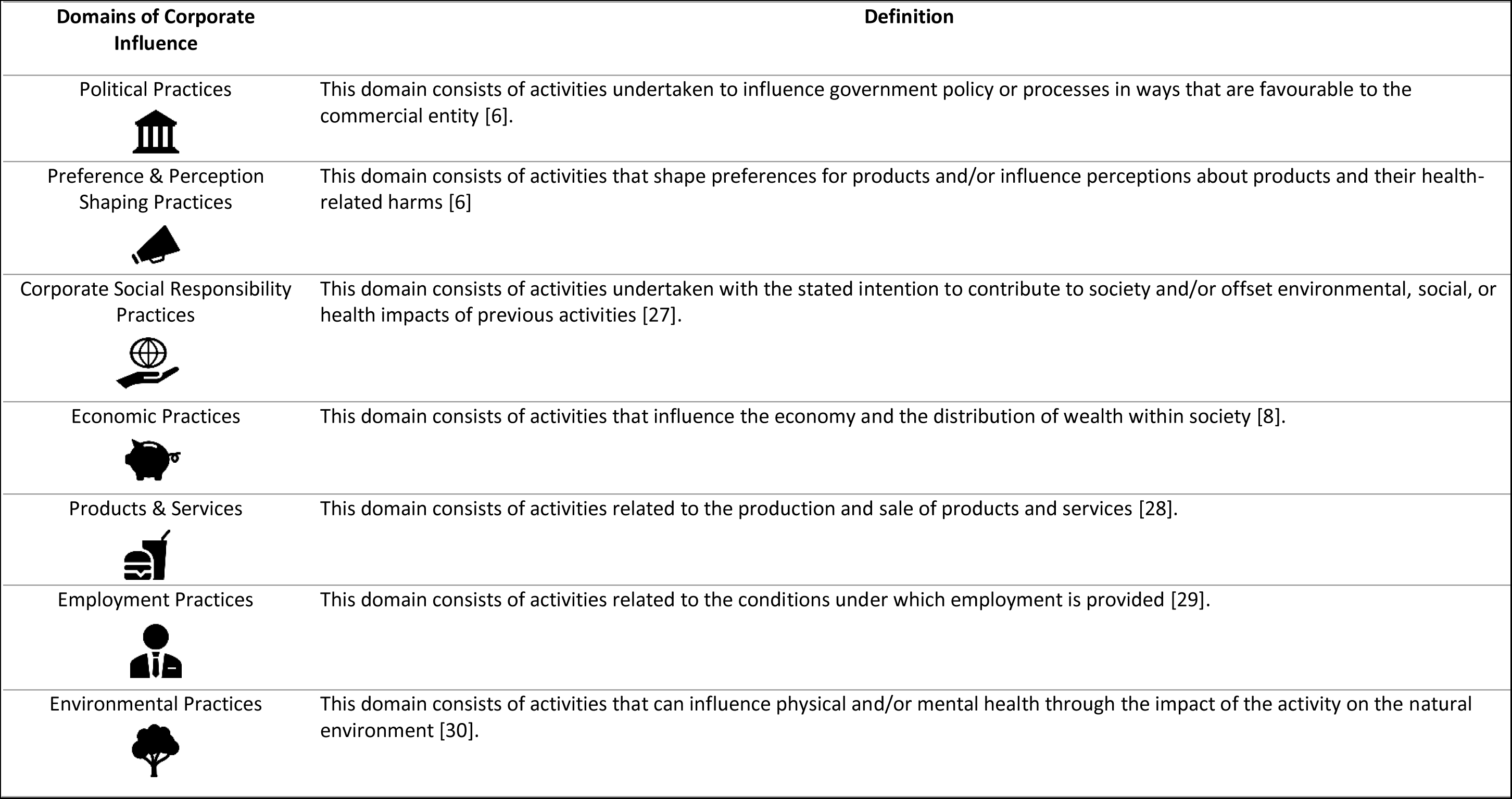
Definitions of Domains of Corporate Influence.

| Domains of Corporate Influence | Definition |
| --- | --- |
| Political Practices | This domain consists of activities undertaken to influence government policy or processes in ways that are favourable to the commercial entity [6]. |
| Preference & Perception Shaping Practices | This domain consists of activities that shape preferences for products and/or influence perceptions about products and their health-related harms [6] |
| Corporate Social Responsibility Practices | This domain consists of activities undertaken with the stated intention to contribute to society and/or offset environmental, social, or health impacts of previous activities [27]. |
| Economic Practices | This domain consists of activities that influence the economy and the distribution of wealth within society [8]. |
| Products & Services | This domain consists of activities related to the production and sale of products and services [28]. |
| Employment Practices | This domain consists of activities related to the conditions under which employment is provided [29]. |
| Environmental Practices | This domain consists of activities that can influence physical and/or mental health through the impact of the activity on the natural environment [30]. |

In Figure 2, we provide a graphical depiction of the domains. We showed that several domains overlap and are interactive (portrayed by positioning the domains in boxes without inter-domain boundaries). For example, some activities (e.g., ‘engage in efforts to influence the public’s perception of health policies’) contain elements of both preference and perception shaping practices and political strategies (overlapping). Other activities (e.g., obscuring conflicts of interest in research) could strengthen activities in other domains (e.g., misrepresenting evidence in policy submissions) (interactive).

**Figure 2.**
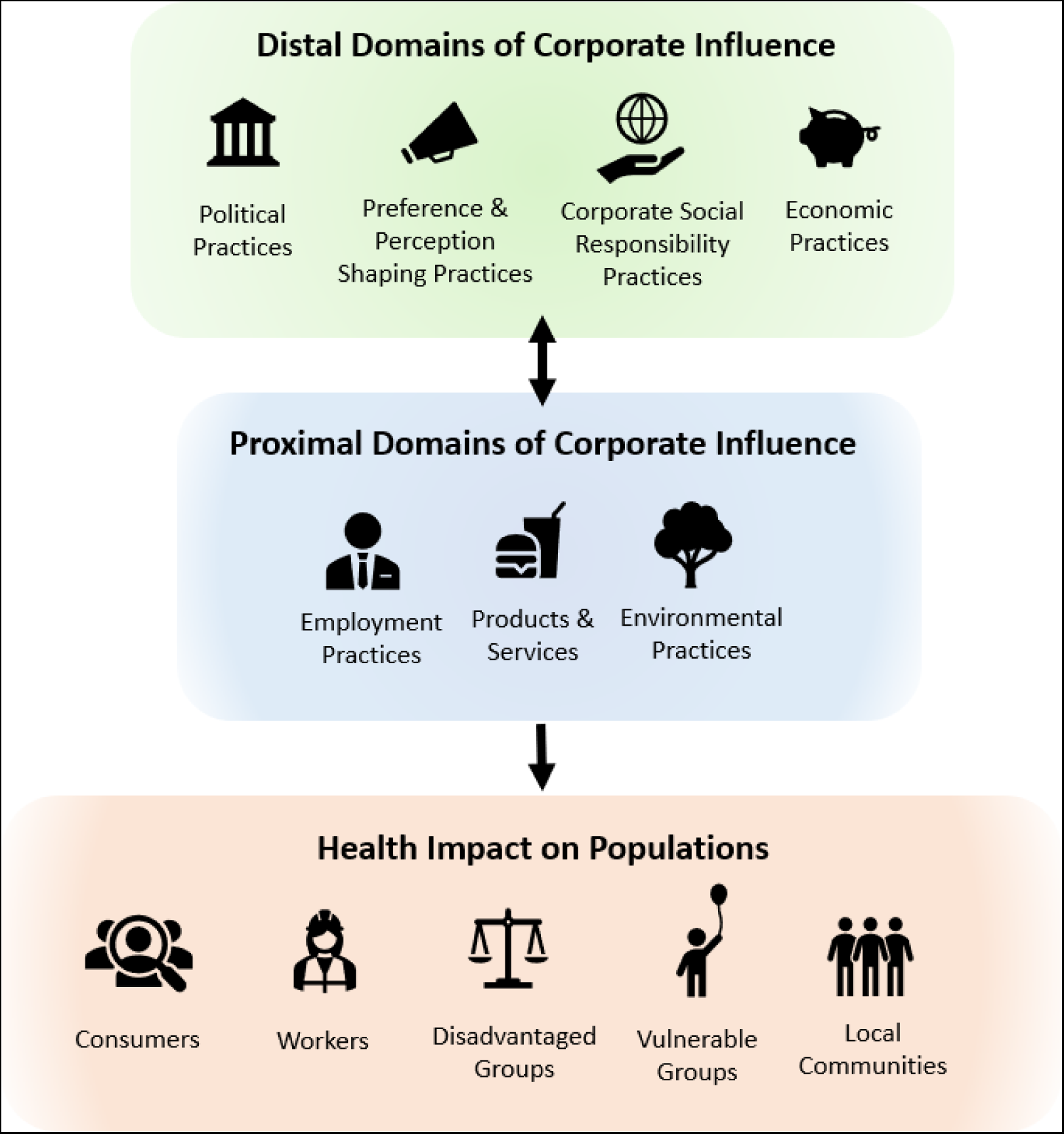
A Model Depicting the Domains of Corporate Influence, their Proximity to Health Outcomes, and the Relevant Population Groups.

Political practices, preference & perception shaping practices, and economic practices are positioned as distal domains (i.e., their impact on population health is indirect). Employment practices, products and services, and environmental practices are positioned as proximal domains (i.e., their impact on population health is direct). A double-headed arrow between the proximal and distal domains suggest that the distal domains enable the proximal domains and the proximal domains reinforce the distal domains. That is, by shaping the political, normative, epistemic, and economic environment in favourable ways, corporations are able to influence the conditions in which they operate (e.g., the environmental impacts for which they are held accountable) [2]. In turn, the proximal domains can simultaneously influence the extent to which commercial entities can engage in the distal domains. For example, transnational companies can leverage their employment of large numbers of people (employment practices) to impose pressure on political and judicial processes (political practices), as was the case with the SNC-Lavalin scandal in Canada in 2019 [25]. Likewise, pharmaceutical companies were able to leverage their production of necessary products to advocate for reduced liability for product-related harms in the United States [26].

Finally, based on our review of the literature and other relevant sources [21,22], we identified five broad population groups that have been consistently studied to assess the impact of corporate activities on population health. These are consumers, workers, disadvantaged groups, vulnerable groups, and local communities located near a production or processing facility (Table 2). These five overlapping groups and the activities that influence their health are described in the section ‘Health Impact on Populations’.

**Table 2.**
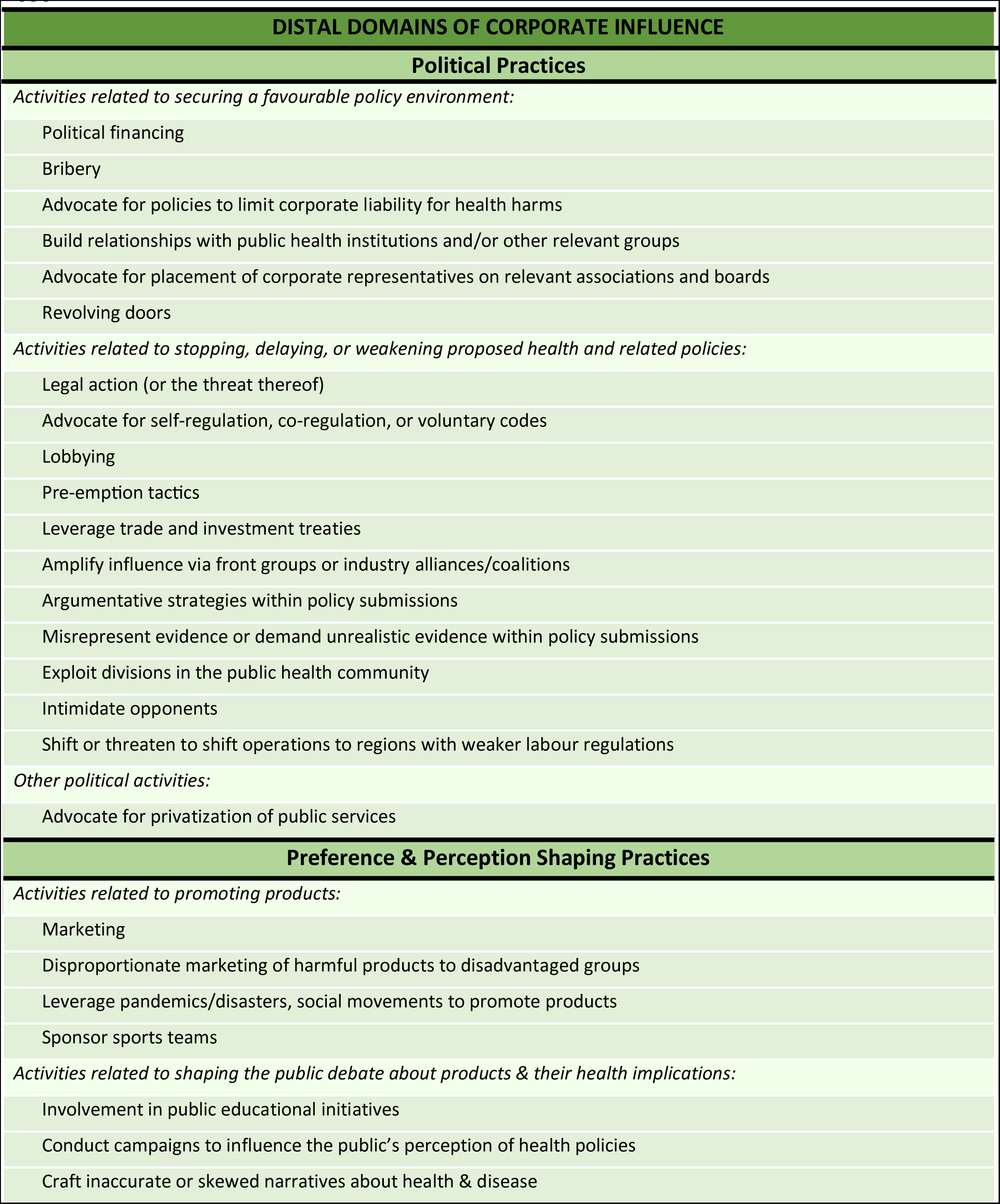
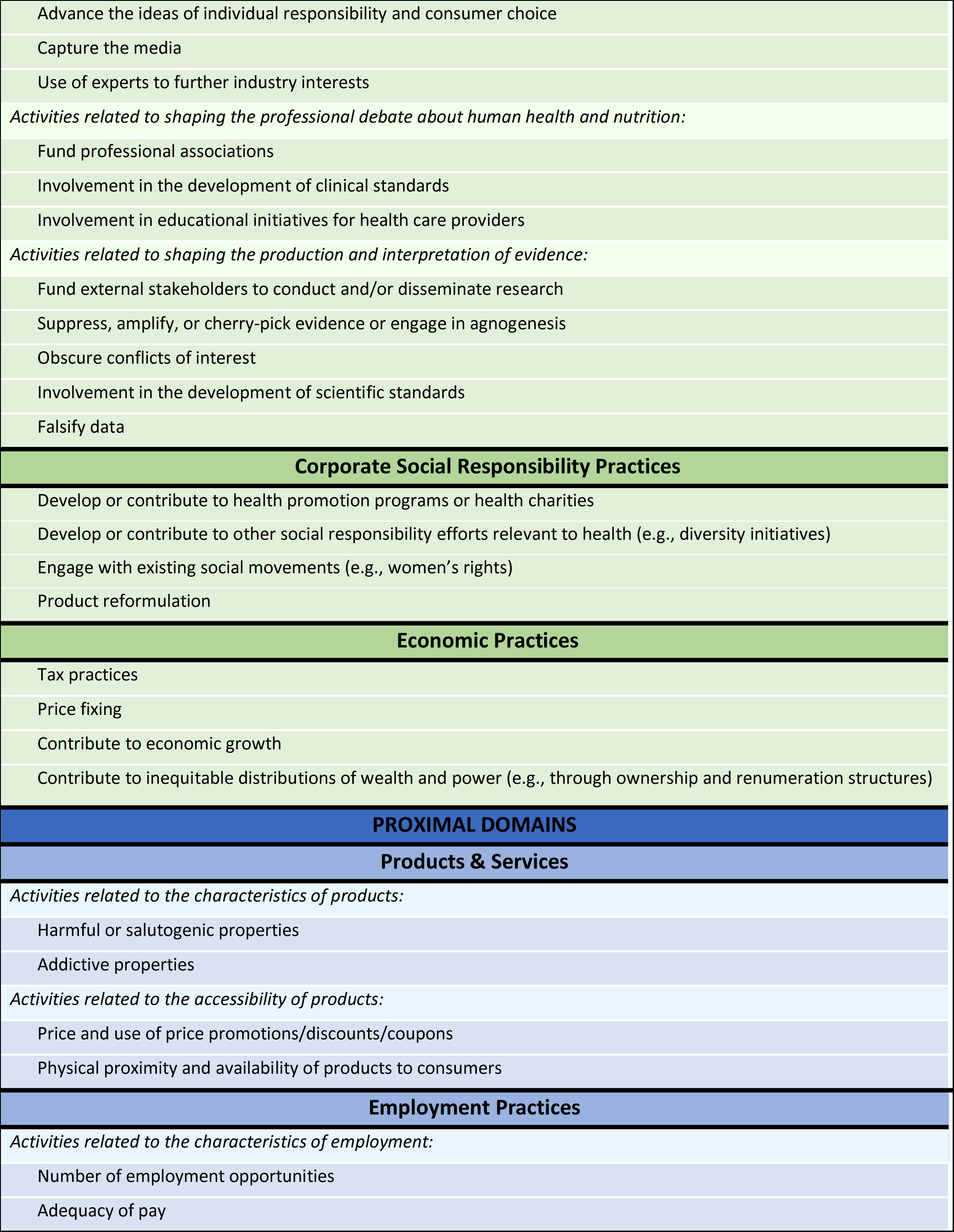
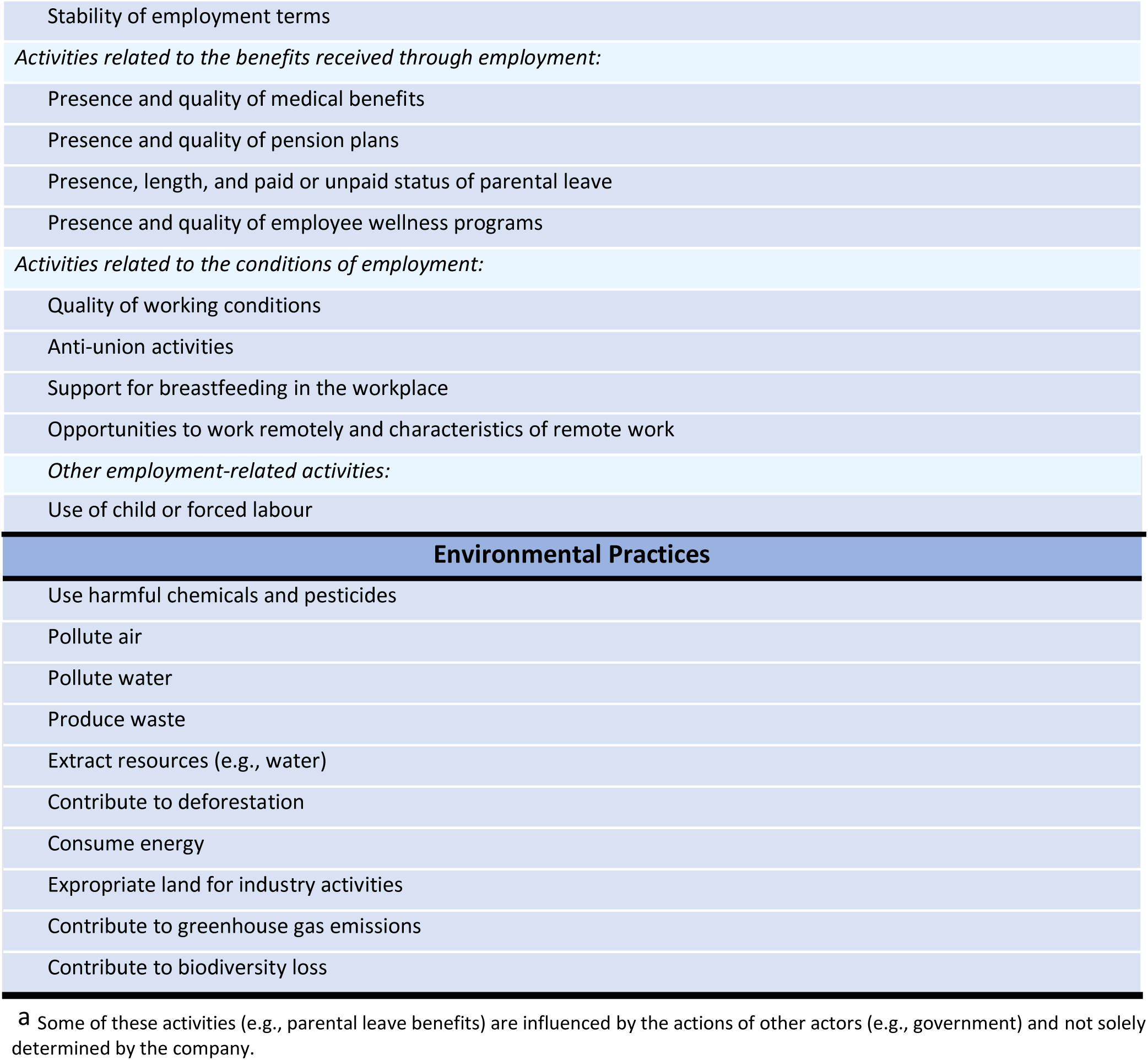
The Corporate Influences on Population Health (HEALTH-CORP) Typology, consisting of seven domains and 70 activities.^a^.

| DISTAL DOMAINS OF CORPORATE INFLUENCE |
| --- |
| Political Practices |
| *Activities related to securing a favourable policy environment:* |
| Political financing |
| Bribery |
| Advocate for policies to limit corporate liability for health harms |
| Build relationships with public health institutions and/or other relevant groups |
| Advocate for placement of corporate representatives on relevant associations and boards |
| Revolving doors |
| *Activities related to stopping, delaying, or weakening proposed health and related policies:* |
| Legal action (or the threat thereof) |
| Advocate for self-regulation, co-regulation, or voluntary codes |
| Lobbying |
| Pre-emption tactics |
| Leverage trade and investment treaties |
| Amplify influence via front groups or industry alliances/coalitions |
| Argumentative strategies within policy submissions |
| Misrepresent evidence or demand unrealistic evidence within policy submissions |
| Exploit divisions in the public health community |
| Intimidate opponents |
| Shift or threaten to shift operations to regions with weaker labour regulations |
| *Other political activities:* |
| Advocate for privatization of public services |
| Preference & Perception Shaping Practices |
| *Activities related to promoting products:* |
| Marketing |
| Disproportionate marketing of harmful products to disadvantaged groups |
| Leverage pandemics/disasters, social movements to promote products |
| Sponsor sports teams |
| *Activities related to shaping the public debate about products & their health implications:* |
| Involvement in public educational initiatives |
| Conduct campaigns to influence the public's perception of health policies |
| Craft inaccurate or skewed narratives about health & disease |
| Advance the ideas of individual responsibility and consumer choice |
| Capture the media |
| Use of experts to further industry interests |
| *Activities related to shaping the professional debate about human health and nutrition:* |
| Fund professional associations |
| Involvement in the development of clinical standards |
| Involvement in educational initiatives for health care providers |
| *Activities related to shaping the production and interpretation of evidence:* |
| Fund external stakeholders to conduct and/or disseminate research |
| Suppress, amplify, or cherry-pick evidence or engage in agnogenesis |
| Obscure conflicts of interest |
| Involvement in the development of scientific standards |
| Falsify data |
| **Corporate Social Responsibility Practices** |
| Develop or contribute to health promotion programs or health charities |
| Develop or contribute to other social responsibility efforts relevant to health (e.g., diversity initiatives) |
| Engage with existing social movements (e.g., women's rights) |
| Product reformulation |
| **Economic Practices** |
| Tax practices |
| Price fixing |
| Contribute to economic growth |
| Contribute to inequitable distributions of wealth and power (e.g., through ownership and remuneration structures) |
| **PROXIMAL DOMAINS** |
| **Products & Services** |
| *Activities related to the characteristics of products:* |
| Harmful or salutogenic properties |
| Addictive properties |
| *Activities related to the accessibility of products:* |
| Price and use of price promotions/discounts/coupons |
| Physical proximity and availability of products to consumers |
| **Employment Practices** |
| *Activities related to the characteristics of employment:* |
| Number of employment opportunities |
| Adequacy of pay |
| Stability of employment terms |
| *Activities related to the benefits received through employment:* |
| Presence and quality of medical benefits |
| Presence and quality of pension plans |
| Presence, length, and paid or unpaid status of parental leave |
| Presence and quality of employee wellness programs |
| *Activities related to the conditions of employment:* |
| Quality of working conditions |
| Anti-union activities |
| Support for breastfeeding in the workplace |
| Opportunities to work remotely and characteristics of remote work |
| *Other employment-related activities:* |
| Use of child or forced labour |
| **Environmental Practices** |
| Use harmful chemicals and pesticides |
| Pollute air |
| Pollute water |
| Produce waste |
| Extract resources (e.g., water) |
| Contribute to deforestation |
| Consume energy |
| Expropriate land for industry activities |
| Contribute to greenhouse gas emissions |
| Contribute to biodiversity loss |

<sup>a</sup>Some of these activities (e.g., parental leave benefits) are influenced by the actions of other actors (e.g., government) and not solely determined by the company.

In the next section, we describe the activities that comprise each domain and report the ways these activities were discussed in the included literature. The number of references pertaining to a specific activity should not be interpreted as an indication of the strength of associated evidence, but rather the number of instances the respective activity was discussed in the included articles. Our descriptions of the activities in the political and preference and perception shaping practices domains are brief as these practices have been discussed in detail elsewhere [3,4,6,7]. The HEALTH-CORP typology is presented in Table 2 (see end of document) and an expanded version with additional details is provided in Additional File 1, Appendix 3.

### Distal Domains of Corporate Influence

#### Political Practices

Corporate involvement in the development and implementation of health policy was a significant focus of many of the included articles and a wide range of corporate strategies were discussed. Corporations were reported to engage in efforts to secure a favourable policy environment, for instance, by contributing to political parties [12,18,20,31–44] and developing relationships with public health institutions [20,27,32,37,45–51]. For example, Maani and colleagues [46] used Freedom of Information Act Requests to access emails between *the Coca Cola Company* and the *Centers for Disease Control and Prevention* (CDC). They demonstrated that *Coca Cola* successfully built a relationship with a senior CDC official, which they leveraged in an attempt to avoid restrictions on sugar-sweetened beverages supported by the *World Health Organization*.

Corporate strategies to stop, delay, or weaken proposed health policies were also frequently discussed. For example, corporations were reported to push for self-regulation schemes [12,18,20,27,31,32,39,49,51–68] and to threaten litigation and/or use existing trade treaties to challenge proposed health legislation [19,67,69–74]. They were reported to engage in a wide breadth of argumentative strategies to challenge proposed health policies. For example, corporations suggested that policies were redundant, conflicting, or misaligned with other regulations or international norms [45,49,52–54,69,75]. Arguments about the impacts of the policy on the economy, employment, equity, and poverty were also common [19,20,31,52–55,63,65,67,71,74,76–80], and economic arguments were considered to be particularly persuasive in low-and-middle income countries (LMICs) [19,67,74]. Many articles also discussed the use of front groups as a way to distance the corporation from its unsavoury activities (e.g., lobbying against health policies) [18,19,32,42,44,48,52,59,63,72,74,81–83].

#### *Preference &* Perception *Shaping Practices*

The ways in which corporations shape preferences for products and influence perceptions about their health-related risks was another major topic of the included articles. Corporations were accused of promoting excessive consumption of harmful products, for example, by engaging in intensive and highly-resourced marketing campaigns that normalize their consumption (e.g., portraying alcohol as part of a normal everyday routine [84]) [12,18,36,42,47,51,63,65,72–74,84–99]. They were reported to influence the public debate about product related-risks by reframing and creating uncertainty about the causes of health issues (e.g., focusing on genetic causes of cancer as opposed to alcohol consumption [100]) [18,20,27,28,31,32,34,36,37,40–42,44,46,48,50–52,56–58,64,65,75,76,79,81,101–104] and acquiring or funding media companies, making it more difficult for public health messages to be heard [20,45,48,49,65,72]. Though commercial entities promoted education as the solution to managing health-related risks [20,45,51,58,66,68,74,80], they were also accused of attempting to shape the public’s understanding of health issues by providing educational resources that promoted their products and/or downplayed the associated health risks (e.g., alcohol [61,62]) [19,20,36,51,63,65,68,80,84,100].

Similarly, commercial actors were accused of promoting products to health professionals by, for example, delivering educational initiatives to health professionals (e.g., the breast-milk substitute (BMS) industry’s funding of medical student retreats [42]) [18–20,49,51,65,80] and influencing the development of clinical standards (e.g., BMS industry’s funding of clinical guidelines for the diagnosis of cows-milk protein allergy [91]) [48,78,91,97,105]. Corporations were also reported to influence the academic debate and the production of science by, for example, by funding universities, think tanks, and scientific conferences, providing scientific awards, developing industry research institutes and suppressing research that is not aligned with their interests (e.g., failing to publish research on health harms of products) [19,20,32,42,46,48,49,51,64,66–68,72,75,80].

#### Corporate Social Responsibility (CSR) Practices

CSR initiatives mentioned in the literature include corporate involvement in health promotion programs and contributions to health charities and causes (e.g., tobacco industry’s funding of HIV initiatives [44]; alcohol industry’s involvement in road safety [37]) [18,19,27,37,39,42,44,51,67,72,91,97,106], some of which had limited evidence of effectiveness [18,27,39,44,72]. Others described CSR initiatives that have implications for health (e.g., diversity, equity, and inclusion efforts) [19,27,39,44,67,68]. Corporations were sometimes reported to exploit existing social movements (e.g., women’s rights [61]) to sell products [27,50,61] or engage in product reformulation (e.g., ‘light’ cigarettes [48]) to suggest that the company is taking action on product-related harms [12,18,36,51,68,92].

Overall, CSR was perceived critically and was seen as being closely related to political practices and preference and perception shaping practices. That is, CSR efforts were seen as attempts to distract from corporate harms [12,18,27,44,48,75,107], shift blame [68,75], enhance brand image and credibility [19,36,44,67,84], influence policymaking [19,37,44,48,68,71], and/or pre-emptively address threats to business practices [55]. In some cases, these strategic uses of CSR were described by corporations in leaked company documents [48]. Millar [62] suggested that ‘good’ corporations engage in CSR efforts genuinely.

#### Economic *Practices*

Externalization of health harms, for example, through tax avoidance [1,36,38,40,41,43,48,62,65,72,86,107–109], was another major topic of the included articles. In this way, corporations were reported to impose health harms onto populations (e.g., chronic disease) without paying for the full cost of these harms. Large monopolies were seen as harmful because they concentrate economic power in one or a few actors, leading to more powerful lobbying efforts [43]. Mendly-Zambo, Raphael, & Taman [106] also discussed the price fixing tactics of *Walmart Canada* and its contributions to food insecurity. Millar [62] described how ‘good’ corporations pay their fair share of taxes and contribute to economic growth.

### Proximal Domain of Corporate Influence

#### Products and Services

Several products (e.g., alcohol, tobacco, and gambling machines) were widely recognized as promoting harm. Many authors discussing products referred to their wide availability and accessibility [36,74,88,98,110,111]. For example, the ubiquity of fast-food outlets and the provision of free BMS in clinics was seen as increasing consumption of these products [36,74,88,110,111]. The features of products were also discussed, such as the hyper-palatable nature of ultra-processed foods [93] and the hyper-engagement features of social media (e.g., endless scroll) [64], which were seen as promoting behaviour some authors described as addictive. Liber [112] discussed some health-promoting products (e.g., vaccines) and suggested that regulation focus on expanding these markets while contracting the markets for health-harming products (e.g., alcohol).

#### *Employment* Practices

Employment practices were not discussed in depth in the included articles. Most articles that discussed employment did so in passing, with reference to health harming practices such as unsafe working conditions, inadequate pay, and limited access to benefits (e.g., parental leave, medical care) [1,12,18,40,41,43,62,65,79,106,113]. Loewenson [113] provided the deepest discussion of employment by describing the global trend towards precarious labour and associated health effects such as social isolation, high blood pressure, and mental ill-health.

Mendly-Zambo, Raphael, and Taman [106] described how *Walmart Canada*’s employment practices (e.g., low wages, anti-union activities) contributed to food insecurity in Canada. In contrast, Millar [62] suggested that ‘good’ businesses create jobs that can have a beneficial effect on human health and described health-promoting activities such as adequate workplace mental health policies.

#### Environmental *Practices*

Health concerns related to environmental practices were not also not a significant topic of the included articles. Activities that were mentioned include chemical and pesticide use, air and water pollution, land clearing/deforestation, ecosystem disruption, consumption of energy and water, production of waste, contributions to greenhouse gases, and contributions to biodiversity loss [1,12,43,62,65,79]. Kadandale, Marten, and Smith [79] described how slash-and-burn land clearing practices used by the palm oil industry created episodes of harmful haze in South-East Asia, which led to thousands of premature deaths and increases in the rates of respiratory, eye, and skin diseases. Montiel [114] described the health harms of deforestation by pointing to reports that land clearing in Indonesia by the palm oil and sugar industries led to the emergence of the Nipah virus through zoonosis (transfer of a virus from animals to humans).

### Health Impact on Populations

Guided by the literature, we describe the domains which are relevant to each of the five identified population groups (Table 3). We also describe specific activities relevant to each group that were discussed within the included literature.

**Table 3.**
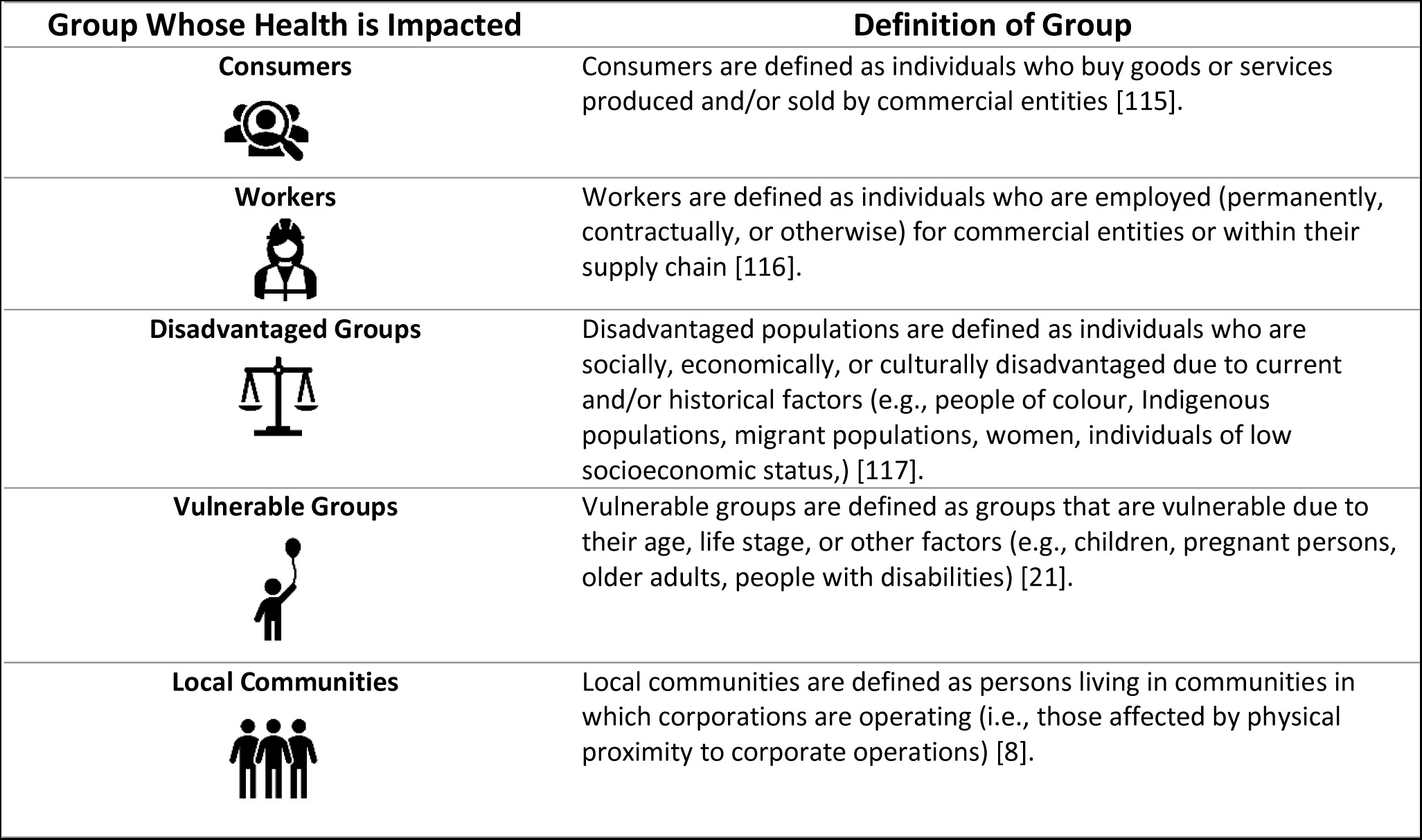
Population Groups Affected by Corporate Activities.

| Group Whose Health is Impacted | Definition of Group |
| --- | --- |
| **Consumers** | Consumers are defined as individuals who buy goods or services produced and/or sold by commercial entities [115]. |
| **Workers** | Workers are defined as individuals who are employed (permanently, contractually, or otherwise) for commercial entities or within their supply chain [116]. |
| **Disadvantaged Groups** | Disadvantaged populations are defined as individuals who are socially, economically, or culturally disadvantaged due to current and/or historical factors (e.g., people of colour, Indigenous populations, migrant populations, women, individuals of low socioeconomic status,) [117]. |
| **Vulnerable Groups** | Vulnerable groups are defined as groups that are vulnerable due to their age, life stage, or other factors (e.g., children, pregnant persons, older adults, people with disabilities) [21]. |
| **Local Communities** | Local communities are defined as persons living in communities in which corporations are operating (i.e., those affected by physical proximity to corporate operations) [8]. |

### Consumers

Consumers, by their nature, are influenced by the properties of consumer goods (products and services). Political and preference and perception shaping practices influence consumers’ health indirectly by creating and shaping the markets for these goods. For example, aggressive marketing can increase the demand for harmful products (e.g., ultra-processed foods) while lobbying and political financing may limit associated protections (e.g., nutrition labelling [49]).

### Workers

The activities influencing the health of workers are, unsurprisingly, those identified in the employment practices domain. These activities shape the opportunities the worker has to obtain optimal health (e.g., access to medical benefits) and their exposure to harmful or salutogenic factors (e.g., pesticides, positive work environments) [118]. Some political practices (e.g., threatening to shift operations to a country with weaker labour standards) may also impact workers (i.e., through placing downward pressure on labour regulations).

### Disadvantaged *Groups*

Disadvantaged groups are more likely to be employed in precarious and unsafe jobs [119] and therefore are likely to be strongly affected by employment practices. These groups may also experience the greatest burden from economic practices such as unfair tax practices that reduce funds for social programs. Environmental practices and preference and perception shaping practices are also relevant to these groups [120,121].

Specific activities described in the literature include disproportionate marketing of unhealthy commodities (e.g., alcohol) to disadvantaged populations [40,43,50,60,72,86,103,110,121–123] and efforts to make unhealthy commodities appealing to women [61,95,98]. Millar described increasing domestic income inequities because of large corporate profits which are distributed amongst the elite, who also lobby for reduced taxation [62]. Some private sectors (e.g., the for-profit prison industry, tobacco industry) were accused of placing undue burdens on certain groups (e.g., Black Americans, Indigenous populations) [1,50,107,124–126]. Corporations were reported to push for privatization, which was seen as widening inequities [62,127]. In some cases, initiatives ostensibly undertaken to improve equity (i.e., efforts to diversify cannabis industry employment) were seen as self-serving attempts to advance industry interests (e.g., increase consumption) [27].

Some articles also described corporate influences on global inequities, including the ‘downward pressure’ on working conditions via corporations’ use of low-wage havens [40,41,113,114], exploitation of the weaker regulatory structures of LMICs, the extraction of wealth from LMICs to HICs [108,114], and the identification of LMICs as ‘emerging markets’ by unhealthy commodity producers (e.g., tobacco) to replace declines in consumption in HICs [36,40,43,72,128].

### Vulnerable *Groups*

The literature described vulnerable groups such as children and pregnant persons being affected by products and associated marketing and educational techniques, as well as the political practices that enable these marketing techniques. Specifically, the included literature discussed food and beverage industry marketing directed towards children and involvement of the industry in schools and other child-centered programming (e.g., distribution of branded school supplies, development of nutrition educational programmes for children) [12,19,43,123]. Pregnant persons and mothers were mostly discussed in the context of the BMS industry and relevant activities included marketing, industry sponsored educational resources, industry interactions with health care professionals, and donations of BMS during emergencies (i.e., ‘crisis marketing’) [18,20,42,47,73,74,91,129]. Other vulnerable groups (e.g., the elderly or people with disabilities) were not frequently discussed.

### *Local* Communities

The health outcomes of local communities are likely be most influenced by environmental and employment practices, especially those occurring in the supply chain, which are enabled by political practices. In the included literature, there was little discussion of specific activities that may influence these communities. One activity that was discussed was the dislocation of local communities [65,113] through, for example, the activities of the extractive industry [113]. This dislocation was reported to lead to social exclusion, poor access to infrastructure, and dependency on mining activities [113].

## Discussion

In this article, we reported on the development of the cross-industry HEALTH-CORP typology, which describes 70 health-relevant corporate activities that are categorized across seven domains of corporate influence. We presented a graphical model that situates these domains in relation to their proximity to health outcomes and identified five population groups whose health is influenced by corporate activities.

These contributions draw from and directly supplement previous work [2–8]. Prior typologies of corporate activities tend to be more narrowly focused on either one particular industry or group of industries [3–5,8,130] and/or a specific type of corporate practice (e.g., political practices) [4,28,131]. To the best of our knowledge, the HEALTH-CORP typology is the most comprehensive typology of health-relevant corporate activities to-date. This novel typology covers a diverse range of corporate activities that were identified based on literature pertaining to 16 different industries. The seven domains we identified are well-aligned with practices proposed by others [2,6,8], indicating agreement on the key corporate practices that require our attention within efforts to mitigate the CDH. The 70 activities identified in the HEALTH-CORP typology add to this work by more comprehensively identifying the array of different activities through which these practices are manifested.

The identification of these activities will support future efforts to measure and monitor corporate activities, a key priority for the CDH field [3,4,8]. Ongoing work by our team includes using the HEALTH-CORP typology to guide the development of a food and beverage (F&B) industry-specific rubric that can be later transformed into an index to measure, monitor, and score individual F&B companies on the diverse set of activities through which they exert influences on health. These efforts, alongside other relevant efforts [21,132,133], will be critical to our ability to track changes in corporate activities over time and to quantitatively assess the impact of corporate activities on specific health outcomes, another priority for the CDH field [24].

The HEALTH-CORP typology may also assist public health professionals, socially minded investors, and civil society in identifying harmful corporate activities that can be targeted for intervention. To achieve meaningful wide-scale change on the CDH, we suggest prioritizing interventions that address distal CDH activities such as the deep integration of corporations into political, academic, and professional structures. For example, implementing regulations that allow industry to fund research but blinding them to the investigators conducting the work [134] may be one measure that could be used to address the power that corporations hold over the production and dissemination of science. By effectively addressing the distal CDH, we may see corollary ‘run-off’ benefits to proximal domains such as products & services (e.g., improved product safety).

Our identification of five population groups whose health is affected by corporate activities may also assist with identifying interventions that can be used to protect the health of specific priority population groups. For example, introducing limits on the number of advertisements for unhealthy commodities on Spanish-language television may help prevent disproportionate marketing of unhealthy goods to Hispanic individuals living in the United States [121]. As suggested by Baum and colleagues [8], the priority placed on different population groups (e.g., local communities versus consumers) and the associated interventions are likely to differ based on the specific characteristics of the particular country or region of interest [8].

Finally, our review and synthesis of a substantial number of CDH articles allowed us to identify key research gaps in the CDH literature. First, the included literature reported extensively on political and preference and perception shaping practices, providing countless examples of how these practices have been manifested in different scenarios. However, some of the proximal domains (i.e., employment practices, environmental practices) were explored only superficially. This gap may be limiting our ability to understand the complex interactions between different types of corporate practices and may also limit our ability to determine which corporate activities are most health-harming and therefore should be prioritized for intervention.

Fortunately, there are robust fields of existing public health research (e.g., environmental health, occupational health) that we can draw on to better understand these activities. Conducting collaborative work with occupational health, environmental health, and CDH scholars may lend fresh insights into these practices, their economic and political drivers, and the associated solutions.

Another gap is the limited investigation of the ways in which commercial entities influence the health of disadvantaged groups, vulnerable groups, and local communities. This may, in part, reflect the relative lack of CDH research on LMICs compared to HICs [reference to related manuscript]. Understanding the impact of corporate practices on these communities will be paramount to addressing domestic and global health inequities. Efforts to advance our understanding of these activities may be facilitated by the Corporate Health Impact Assessment tool developed by Baum and colleagues [8], which can be used to conduct in-depth investigations of corporate practices within particular regions of interest.

Researchers can also expand our understanding of the role of specific actors by conducting community-based participatory research [135] with communities of interest to better understand the corporate activities that most affect them and determine how individuals in these communities weigh positive impacts of corporate involvement in their community (e.g., employment) against negative impacts (e.g., water pollution).

## Limitations

We were able to review and synthesize 116 articles on the CDH across 16 industries to develop a fairly comprehensive typology of corporate activities that can influence health. We believe the HEALTH-CORP typology will be useful for advancing scholarship and practice on the CDH. However, the typology nor our associated model can completely explain the complex system through which commercial entities influence health. Moreover, other domains of corporate influence, such as corporate activities that exert influence over patterns of conflict and migration, may continue to emerge as CDH research advances.

We have focused on corporate actions in this article, rather than the systems and structures (e.g., neoliberalism) that drive these actions, or the influence of other actors (e.g., government) in co-creating these actions (e.g., privatization of services). The benefits of this approach are that actions are observable and observing them may lend insight into the associated structures. We included all activities that could be reasonably linked to human health; however, it was beyond the scope of this review to assess the strength of evidence of the relationship of each activity to health.

Moreover, most of the included literature described corporate practices from a negative perspective that seemed to suggest that corporate actions are driven by profit-seeking and sometimes a willful neglect of the associated harms to health. In many cases (e.g., tobacco industry’s attempts to sow doubt about the health harms of cigarettes), this perception may be accurate whereas in other cases (e.g., aggressive marketing) corporations may be driven by other factors inherent to our economic system (e.g., fiduciary duty to maximize profits and revenues). The proposed HEALTH-CORP model and typology (and CDH literature in general) may benefit from deeper consideration of the benefits some large companies provide to society that the current economic system facilitates (e.g., inexpensive goods manufactured efficiently at large-scales).

Our eligibility criteria did not include grey and non-English literature which may have improved the comprehensiveness of the HEALTH-CORP typology. Importantly, we also limited our review to articles that directly engaged with CDH terms, thereby excluding research that may describe health-relevant corporate practices without engaging with CDH concepts. We made this decision for feasibility purposes and because this strategy allowed us to identify key gaps in CDH literature, a contribution of this review.

## Conclusions

Scholarship on the CDH has documented the wide-ranging influence that corporate activities can have on population health. In this article, we make three contributions to the CDH literature. First, we developed the HEALTH-CORP typology, which describes 70 corporate activities that can influence population health across industries and categorizes these activities into seven domains of corporate influence. Second, we situate the domains based on their proximity to health outcomes and identify five population groups to consider when evaluating the health impact of corporate practices. Finally, we leverage our findings to reveal key gaps in CDH literature and recommend future avenues for CDH research. We believe these contributions will be useful for advancing public health research and practice related to the CDH.

## Data Availability

The data for this study are published academic articles which are available from the respective publishers (see Supplementary Material, Appendix 2 for the characteristics of included articles). In addition, we uploaded the following files to Open Science Framework (DOI 10.17605/OSF.IO/TG9S7) to support data availability: 1) a .csv file containing a list of the articles that underwent title and abstract screening in our study and the respective screening decisions that were assigned, and 2) .ris files containing the citations to the respective articles and the assigned screening decisions, which can be uploaded into a reference manager. Interested parties can contact the corresponding author for additional information.

https://osf.io/tg9s7/

## List of abbreviations

CDH: Commercial determinants of health SDH: Social determinants of health
HEALTH-CORP: Corporate Influences on Population Health PRISMA-ScR: PRISMA Extension for Scoping Reviews
CSR: Corporate Social Responsibility
CDC: Centers for Disease Control and Prevention HICs: High-income countries
LMICs: Low- and middle-income countries F&B: Food and beverage

## Declarations

### Ethical approval and consent to participate

Not applicable.

### Consent for publication

Not applicable.

### Availability of data and materials

The data for this study are published academic articles which are available from the respective publishers (see Additional File 1, Appendix 2 for the characteristics of included articles). In addition, we uploaded the following files to Open Science Framework (DOI 10.17605/OSF.IO/TG9S7) to support data availability: 1) a .csv file containing a list of the articles that underwent title and abstract screening in our study and the respective screening decisions that were assigned, and 2) .ris files containing the citations to the respective articles and the assigned screening decisions, which can be uploaded into a reference manager. Interested parties can contact the corresponding author for additional information.

### Authors’ contributions

Burgess conceptualized the study, completed the search and review process, synthesized the findings, and led the writing of the manuscript. K. Nyhan guided the review methodology and search strategy and contributed to the conceptualization of the study and revision of the manuscript. N. Freudenberg contributed to the intellectual content and revision of the manuscript and the development and revision of the typology and model. Y. Ransome supervised the research and contributed to the conceptualization of the study and revision of the manuscript.

## Acknowledgements

We would like to acknowledge the CDH experts who provided input on the HEALTH-CORP typology in the context of a related project. We would also like to acknowledge Dr. Trace Kershaw, Dr. Ijeoma Opara, and Dr. Rafael Perez-Escamilla for the useful feedback they provided related to this review.

## References

1. Freudenberg N, Lee K, Buse K, Collin J, Crosbie E, Friel S, et al. Defining Priorities for Action and Research on the Commercial Determinants of Health: A Conceptual Review. Am J Public Health [Internet]. 2021;111:2202–11. Available from: http://ovidsp.ovid.com/ovidweb.cgi?T=JS&PAGE=reference&D=emexa&NEWS=N&AN=636863634

2. Gilmore AB, Fabbri A, Baum F, Bertscher A, Bondy K, Chang H-J, et al. Defining and conceptualising the commercial determinants of health. Lancet [Internet]. 2023 [cited 2023 Apr 20];401:1194–213. Available from: http://www.ncbi.nlm.nih.gov/pubmed/36966782

3. Bennett E, Topp S, Moodie A. National Public Health Surveillance of Corporations in Key Unhealthy Commodity Industries–a scoping review and framework synthesis. Int J Health Policy Manag. 2023;1–41.

4. Mialon M, Swinburn B, Sacks G. A proposed approach to systematically identify and monitor the corporate political activity of the food industry with respect to public health using publicly available information. Obesity Reviews [Internet]. 2015 [cited 2022 Feb 2];16:519–30. Available from: https://onlinelibrary.wiley.com/doi/full/10.1111/obr.12289

5. Ulucanlar S, Fooks GJ, Gilmore AB. The Policy Dystopia Model: An Interpretive Analysis of Tobacco Industry Political Activity. PLoS Med [Internet]. 2016 [cited 2021 Sep 16];13:e1002125. Available from: http://medicine.plosjournals.org/perlserv/?request=index-html&issn=1549-1676

6. Madureira Lima J, Galea S. Corporate practices and health: A framework and mechanisms. Global Health [Internet]. 2018 [cited 2021 May 9];14:1–12. Available from: https://www.scopus.com/inward/record.uri?eid=2-s2.0-85042139498&doi=10.1186%2Fs12992-018-0336-y&partnerID=40&md5=737e6e39d95d958e17845d3589cebe05

7. Ulucanlar S, Lauber K, Fabbri A, Hawkins B, Mialon M, Hancock L, et al. Corporate Political Activity: Taxonomies and Model of Corporate Influence on Public Policy. 2023 [cited 2023 Apr 30]; Available from: 10.34172/ijhpm.2023.7292

8. Baum FE, Sanders DM, Fisher M, Anaf J, Freudenberg N, Friel S, et al. Assessing the health impact of transnational corporations: Its importance and a framework. Global Health [Internet]. 2016 [cited 2021 Mar 9];12:1–7. Available from: https://link.springer.com/articles/10.1186/s12992-016-0164-x

9. Munn Z, Peters MDJ, Stern C, Tufanaru C, McArthur A, Aromataris E. Systematic review or scoping review? Guidance for authors when choosing between a systematic or scoping review approach. BMC Med Res Methodol [Internet]. 2018 [cited 2023 Apr 13];18:1–7. Available from: https://bmcmedresmethodol.biomedcentral.com/articles/10.1186/s12874-018-0611-x

10. Bearman M, Dawson P. Qualitative synthesis and systematic review in health professions education. Med Educ [Internet]. 2013 [cited 2023 Dec 7];47:252–60. Available from: https://onlinelibrary.wiley.com/doi/full/10.1111/medu.12092

11. Tricco AC, Lillie E, Zarin W, O’Brien KK, Colquhoun H, Levac D, et al. PRISMA extension for scoping reviews (PRISMA-ScR): Checklist and explanation. Ann Intern Med [Internet]. 2018 [cited 2023 Mar 12];169:467–73. Available from: https://www.acpjournals.org/doi/10.7326/M18-0850

12. De Lacy-Vawdon C, Livingstone C. Defining the commercial determinants of health: A systematic review. BMC Public Health [Internet]. 2020 [cited 2021 May 15];20:1022. Available from: https://rd.springer.com/article/10.1186/s12889-020-09126-1

13. Mendeley Desktop. Elselvier;

14. Ouzzani M, Hammady H, Fedorowicz Z, Elmagarmid A. Rayyan - a web and mobile app for systematic reviews. Syst Rev. 2016;5.

15. The EndNote Team. EndNote. Philadelphia, PA: Clarivate; 2013.

16. QSR International. NVivo 12 qualitative data analysis software. 2020.

17. Glaser BG. The Constant Comparative Method of Qualitative Analysis. Soc Probl. 1965;12:436–45.

18. Baker P, Russ K, Kang M, Santos TM, Neves PAR, Smith J, et al. Globalization, first-foods systems transformations and corporate power: a synthesis of literature and data on the market and political practices of the transnational baby food industry. Global Health [Internet]. 2021;17:58. Available from: https://www.scopus.com/inward/record.uri?eid=2-s2.0-85106677038&doi=10.1186%2Fs12992-021-00708-1&partnerID=40&md5=647acb4b9973cc4e440bcece840b947c

19. Mialon M, Gaitan Charry DA, Cediel G, Crosbie E, Baeza Scagliusi F, Pérez Tamayo EM. “the architecture of the state was transformed in favour of the interests of companies”: Corporate political activity of the food industry in Colombia. Global Health [Internet]. 2020;16:97. Available from: http://www.globalizationandhealth.com/

20. Tanrikulu H, Neri D, Robertson A, Mialon M. Corporate political activity of the baby food industry: The example of Nestlé in the United States of America. Int Breastfeed J [Internet]. 2020;15:22. Available from: https://www.scopus.com/inward/record.uri?eid=2-s2.0-85083098748&doi=10.1186%2Fs13006-020-00268-x&partnerID=40&md5=a89f6a235b94c64a067293a460d9e3d6

21. Global Access to Nutrition Index 2021: Methodology. 2020.

22. Improving people’s health [Internet]. ShareAction. [cited 2023 Nov 4]. Available from: https://shareaction.org/global-issues/better-health

23. Peters M, Godfrey C, McInerney P, Munn Z, Tricco A, Khalil H. Chapter 11: Scoping Reviews. In: Aromataris E, Munn Z, editors. JBI Manual for Evidence Synthesis. JBI; 2020.

24. Burgess R, Nyhan K, Dharia N, Freudenberg N, Ransome Y. Characteristics of commercial determinants of health research on corporate activities: A scoping review. Provisionally accepted at PLOS One. 2024;

25. Gollom M. What you need to know about the SNC-Lavalin affair. CBC News. 2019 Feb 13;

26. Holland MS. Liability for Vaccine Injury: the United States, the European Union, and the Developing World. Emory Law J [Internet]. 2017;67:415–62. Available from: http://eds.a.ebscohost.com/eds/detail/detail?vid=0&sid=de62705b-3bfe-4ca8-86e0-5eed44ad3cb8%40sdc-v-sessmgr02&bdata=JnNpdGU9ZWRzLWxpdmU%3D#AN=129043427&db=aph

27. Wakefield T, Glantz SA, Apollonio DE, T. W, S.A. G. Content Analysis of the Corporate Social Responsibility Practices of 9 Major Cannabis Companies in Canada and the US. JAMA Netw Open [Internet]. 2022;E2228088. Available from: https://jamanetwork.com/journals/jamanetworkopen

28. Knai C, Petticrew M, Capewell S, Cassidy R, Collin J, Cummins S, et al. The case for developing a cohesive systems approach to research across unhealthy commodity industries. BMJ Glob Health [Internet]. 2021;6:e003543. Available from: https://gh.bmj.com/

29. Occupational Health [Internet]. International Labour Organization. [cited 2023 Feb 28]. Available from: https://www.ilo.org/safework/areasofwork/occupational-health/lang--en/index.htm

30. Sattler B. Environmental Health. Policy Polit Nurs Pract. 2003;4:4–5.

31. Mialon M. An overview of the commercial determinants of health. Global Health. BioMed Central Ltd; 2020.

32. Buse K, Tanaka S, Hawkes S. Healthy people and healthy profits? Elaborating a conceptual framework for governing the commercial determinants of non-communicable diseases and identifying options for reducing risk exposure. Global Health. BioMed Central Ltd.; 2017.

33. Brown T. Legislative Capture: Critical Considerations in the Commercial Determinants of Health. SSRN Electronic Journal [Internet]. 2019; Available from: https://www.ssrn.com/abstract=3687855

34. van Schalkwyk MCI, Petticrew M, Cassidy R, Adams P, McKee M, Reynolds J, et al. A public health approach to gambling regulation: countering powerful influences. Lancet Public Health. 2021;6:e614–9.

35. Wiist WH. The Foundations of Corporate Strategies Comment on “‘Part of the Solution’: Food Corporation Strategies for Regulatory Capture and Legitimacy”. Int J Health Policy Manag [Internet]. 2022; Available from: http://ovidsp.ovid.com/ovidweb.cgi?T=JS&PAGE=reference&D=medp&NEWS=N&AN=35397485

36. Chavez-Ugalde Y, Jago R, Toumpakari Z, Egan M, Cummins S, White M, et al. Conceptualizing the commercial determinants of dietary behaviors associated with obesity: A systematic review using principles from critical interpretative synthesis. Obes Sci Pract [Internet]. 2021;7:473–86. Available from: https://www.scopus.com/inward/record.uri?eid=2-s2.0-85103578531&doi=10.1002%2Fosp4.507&partnerID=40&md5=fbbb0335599e89a0add6892a7ec20c05

37. Hoe C, Taber N, Champagne S, Bachani AM. Drink, but don’t drive? The alcohol industry’s involvement in global road safety. Health Policy Plan [Internet]. 2020;35:1328–38. Available from: http://ovidsp.ovid.com/ovidweb.cgi?T=JS&PAGE=reference&D=emed22&NEWS=N&AN=633507007

38. Wiist WH. Mechanisms Underlying Corporations as Determinants of Health. Am J Public Health [Internet]. 2019 [cited 2022 Jan 3];109:e1. Available from: http://ovidsp.ovid.com/ovidweb.cgi?T=JS&PAGE=reference&D=emed20&NEWS=N&AN=625989148

39. Eastmure E, Cummins S, Sparks L. Non-market strategy as a framework for exploring commercial involvement in health policy: A primer. Soc Sci Med [Internet]. 2020;262:113257. Available from: https://www.scopus.com/inward/record.uri?eid=2-s2.0-85089080870&doi=10.1016%2Fj.socscimed.2020.113257&partnerID=40&md5=d263d4fad0fb157d521ed1b9ba25fc53

40. Jamieson L, Gibson B, Thomson WM, L. J, B. G. Oral health inequalities and the corporate determinants of health: A commentary. Int J Environ Res Public Health [Internet]. 2020;17:1–6. Available from: https://www.mdpi.com/1660-4601/17/18/6529/pdf

41. McKee M, Stuckler D, M. M, McKee M, Stuckler D. Revisiting the corporate and commercial determinants of health. Am J Public Health [Internet]. 2018 [cited 2022 Jan 3];108:1167–70. Available from: https://www.scopus.com/inward/record.uri?eid=2-s2.0-85051184487&doi=10.2105%2FAJPH.2018.304510&partnerID=40&md5=407ebdbc80f5eeb253021388f017af12

42. Cossez E, Baker P, Mialon M. ‘The second mother’: How the baby food industry captures science, health professions and civil society in France. Matern Child Nutr [Internet]. 2022;18:e13301. Available from: http://onlinelibrary.wiley.com/journal/10.1111/(ISSN)1740-8709

43. Freudenberg N. Responding to Food Industry Initiatives to Be “Part of the Solution” Comment on “‘Part of the Solution’: Food Corporation Strategies for Regulatory Capture and Legitimacy.” Int J Health Policy Manag [Internet]. 2022;11:2740–3. Available from: http://ovidsp.ovid.com/ovidweb.cgi?T=JS&PAGE=reference&D=emexb&NEWS=N&AN=638690

44. Adams PJJ, Rychert M, Wilkins C. Policy influence and the legalized cannabis industry: learnings from other addictive consumption industries. Addiction [Internet]. 2021;116:2939–46. Available from: https://www.scopus.com/inward/record.uri?eid=2-s2.0-85103170796&doi=10.1111%2Fadd.15483&partnerID=40&md5=b9e5eb3fa6837fad7c7b1da8f850f427

45. Kroker-Lobos MF, Morales LA, Ramirez-Zea M, Vandevijvere S, Champagne B, Mialon M, et al. Two countries, similar practices: The political practices of the food industry influencing the adoption of key public health nutrition policies in Guatemala and Panama. Public Health Nutr [Internet]. 2022;1–33. Available from: http://ovidsp.ovid.com/ovidweb.cgi?T=JS&PAGE=reference&D=emexb&NEWS=N&AN=638801438

46. Maani Hessari N, Ruskin G, McKee M, Stuckler D. Public Meets Private: Conversations Between Coca-Cola and the CDC. Milbank Quarterly [Internet]. 2019;97:74–90. Available from: http://onlinelibrary.wiley.com/journal/10.1111/(ISSN)1468-0009

47. Hastings G, Angus K, Eadie D, Hunt K. Selling second best: How infant formula marketing works. Global Health [Internet]. 2020;16:77. Available from: http://ovidsp.ovid.com/ovidweb.cgi?T=JS&PAGE=reference&D=emed21&NEWS=N&AN=632727368

48. Hird TR, Gallagher AWA, Evans-Reeves K, Zatonski M, Dance S, Diethelm PA, et al. Understanding the long-term policy influence strategies of the tobacco industry: two contemporary case studies. Tob Control [Internet]. 2022;31:297–307. Available from: https://www.scopus.com/inward/record.uri?eid=2-s2.0-85125690415&doi=10.1136%2Ftobaccocontrol-2021-057030&partnerID=40&md5=39c263575d3e9fdd09f112b4e324b535

49. Mialon M, Charry DAG, Cediel G, Crosbie E, Scagliusi FB, Tamayo EMP. “I had never seen so many lobbyists”: Food industry political practices during the development of a new nutrition front-of-pack labelling system in Colombia. Public Health Nutr [Internet]. 2021;24:2737–45. Available from: http://ovidsp.ovid.com/ovidweb.cgi?T=JS&PAGE=reference&D=emed22&NEWS=N&AN=632668742

50. Maani N, Van Schalkwyk MC, Petticrew M, Galea S. The Commercial Determinants of Three Contemporary National Crises: How Corporate Practices Intersect With the COVID-19 Pandemic, Economic Downturn, and Racial Inequity. Milbank Quarterly [Internet]. 2021 [cited 2022 Feb 2];99:503–18. Available from: https://www.scopus.com/inward/record.uri?eid=2-s2.0-85103384553&doi=10.1111%2F1468-0009.12510&partnerID=40&md5=0cb90796e1918e0b9a4cc9becfd5fa48

51. Mialon M, Crosbie E, Sacks G, M. M, E. C, Sacks G. AO - Mialon MO https://orcid.org/0000-0002-9883-6441. Mapping of food industry strategies to influence public health policy, research and practice in South Africa. Int J Public Health [Internet]. 2020;65:1027–36. Available from: http://ovidsp.ovid.com/ovidweb.cgi?T=JS&PAGE=reference&D=emed21&NEWS=N&AN=632488017

52. Lauber K, Hunt D, Gilmore AB, Rutter H. Corporate political activity in the context of unhealthy food advertising restrictions across Transport for London: A qualitative case study. PLoS Med. 2021;18.

53. Hunt D. How food companies use social media to influence policy debates: A framework of Australian ultra-processed food industry Twitter data. Public Health Nutr. 2021;24:3124–35.

54. Lauber K, Ralston R, Mialon M, Carriedo A, Gilmore AB. Non-communicable disease governance in the era of the sustainable development goals: A qualitative analysis of food industry framing in WHO consultations. Global Health. 2020;16.

55. Lacy-Nichols J, Marten R. Power and the commercial determinants of health: Ideas for a research agenda. BMJ Glob Health. BMJ Publishing Group; 2021.

56. Knai C, Petticrew M, Mays N, Capewell S, Cassidy R, Cummins S, et al. Systems Thinking as a Framework for Analyzing Commercial Determinants of Health. Milbank Quarterly. 2018;96:472– 98.

57. Campbell N, Browne S, Claudy M, Mialon M, Hercberg S, Goiana-da-Silva F, et al. The Gift of Data: Industry-Led Food Reformulation and the Obesity Crisis in Europe. Journal of Public Policy and Marketing. 2021;40:389–402.

58. Lacy-Nichols J, Scrinis G, Carey R. The politics of voluntary self-regulation: Insights from the development and promotion of the Australian Beverages Council’s Commitment. Public Health Nutr. 2020;23:564–75.

59. Backholer K, Baum F, Finlay SM, Friel S, Giles-Corti B, Jones A, et al. Australia in 2030: what is our path to health for all? Medical Journal of Australia. 2021;214:S5–40.

60. S.E. H, P. J, R.T. N, Collin J. AO - Hill Rima T.; ORCID: https://orcid.org/0000-0001-8800-5591 SE; O https://orcid.org/0000-0003-3555-433X AO-N, Hill SE, Johns P, et al. From silos to policy coherence: tobacco control, unhealthy commodity industries and the commercial determinants of health. Tob Control [Internet]. 2022;31:322–7. Available from: https://www.scopus.com/inward/record.uri?eid=2-s2.0-85125691950&doi=10.1136%2Ftobaccocontrol-2021-057136&partnerID=40&md5=eaf9dd75ae1ce88ae3e426ec832340ba

61. Hill SE, Friel S. ‘As long as it comes off as a cigarette ad, not a civil rights message’: Gender, inequality and the commercial determinants of health. Int J Environ Res Public Health [Internet]. 2020;17:1–19. Available from: https://www.mdpi.com/1660-4601/17/21/7902/pdf

62. Millar JS. The corporate determinants of health: How big business affects our health, and the need for government action! Canadian Journal of Public Health [Internet]. 2013 [cited 2021 May 11];104:e327–9. Available from: http://journal.cpha.ca/index.php/cjph/article/download/3849/2840

63. Van Schalkwyk MC, Maani N, Cohen J, Mckee M, Petticrew M. Our Postpandemic World: What Will It Take to Build a Better Future for People and Planet? Milbank Quarterly [Internet]. 2021;99:467–502. Available from: https://www.scopus.com/inward/record.uri?eid=2-s2.0-85103403620&doi=10.1111%2F1468-0009.12508&partnerID=40&md5=1d3dd9e1dc9d712ab17e510f3e23d636

64. Zenone M, Kenworthy N, Maani N. The Social Media Industry as a Commercial Determinant of Health. Int J Health Policy Manag [Internet]. 2023;12. Available from: http://ovidsp.ovid.com/ovidweb.cgi?T=JS&PAGE=reference&D=medp&NEWS=N&AN=35490262

65. Wood B, Baker P, Sacks G. Conceptualising the Commercial Determinants of Health Using a Power Lens: A Review and Synthesis of Existing Frameworks. Int J Health Policy Manag [Internet]. 2022;11:1251–61. Available from: http://ovidsp.ovid.com/ovidweb.cgi?T=JS&PAGE=reference&D=emexb&NEWS=N&AN=634395313

66. Brown T. Public Health versus Alcohol Industry Compliance Laws: A Case of Industry Capture? J Law Med [Internet]. 2020;27:1047–73. Available from: https://www.scopus.com/inward/record.uri?eid=2-s2.0-85090260756&partnerID=40&md5=2b8d09a793fa746a935cd0f17d30a7c3

67. Abdool Karim S, Kruger P, Hofman K. Industry strategies in the parliamentary process of adopting a sugar-sweetened beverage tax in South Africa: a systematic mapping. Global Health [Internet]. 2020;16:116. Available from: http://www.globalizationandhealth.com/

68. Mialon M, Mialon J, Calixto Andrade G, Jean-Claude M. ‘We must have a sufficient level of profitability’: food industry submissions to the French parliamentary inquiry on industrial food. Crit Public Health [Internet]. 2020;30:457–67. Available from: https://www.tandf.co.uk/journals/titles/09581596.asp

69. Mialon M, Khandpur N, Laís MA, Martins APB. Arguments used by trade associations during the early development of a new front-of-pack nutrition labelling system in Brazil. Public Health Nutr. 2020;

70. Barlow P, Thow AM, P. B. Neoliberal discourse, actor power, and the politics of nutrition policy: A qualitative analysis of informal challenges to nutrition labelling regulations at the World Trade Organization, 2007-2019. Soc Sci Med [Internet]. 2021;273:113761. Available from: https://www.elsevier.com/locate/socscimed

71. Clare K, Maani N, Milner J. Meat, money and messaging: How the environmental and health harms of red and processed meat consumption are framed by the meat industry. Food Policy [Internet]. 2022;109. Available from: https://www.scopus.com/inward/record.uri?eid=2-s2.0-85127324192&doi=10.1016%2Fj.foodpol.2022.102234&partnerID=40&md5=05065edaeda523067beeab20336ba5af

72. Hoe C, Weiger C, Minosa MKR, Alonso F, Koon AD, Cohen JE. Strategies to expand corporate autonomy by the tobacco, alcohol and sugar-sweetened beverage industry: a scoping review of reviews. Global Health [Internet]. 2022;18:17. Available from: http://www.globalizationandhealth.com/

73. Russ K, Baker P, Byrd M, Kang M, Siregar RN, Zahid H, et al. What You Don’t Know About the Codex Can Hurt You: How Trade Policy Trumps Global Health Governance in Infant and Young Child Nutrition. Int J Health Policy Manag [Internet]. 2021;10:983–97. Available from: http://ovidsp.ovid.com/ovidweb.cgi?T=JS&PAGE=reference&D=medp&NEWS=N&AN=34634868

74. Baker P, Zambrano P, Mathisen R, Singh-Vergeire MRR, Escober AEE, Mialon M, et al. Breastfeeding, first-food systems and corporate power: a case study on the market and political practices of the transnational baby food industry and public health resistance in the Philippines. Global Health [Internet]. 2021;17. Available from: https://www.scopus.com/inward/record.uri?eid=2-s2.0-85118243807&doi=10.1186%2Fs12992-021-00774-5&partnerID=40&md5=1844e47474ba77e1e2b1c38668ddc4ff

75. Brisbois B, Feagan M, Stime B, Paz IK, Berbés-Blázquez M, Gaibor J, et al. Mining, Colonial Legacies, and Neoliberalism: A Political Ecology of Health Knowledge: Minerıa, legados coloniales y neoliberalismo: una ecologıa polıtica del conocimiento en salud. New Solutions [Internet]. 2021;31:48–64. Available from: https://www.scopus.com/inward/record.uri?eid=2-s2.0-85102708084&doi=10.1177%2f10482911211001051&partnerID=40&md5=3342652af1e49caa84d5a3a9a88c62e5

76. Mialon M, Crosbie E, Sacks G. Mapping of food industry strategies to influence public health policy, research and practice in South Africa. Int J Public Health. 2020;65:1027–36.

77. McHardy J. The WHO FCTC’s lessons for addressing the commercial determinants of health. Health Promot Int. 2021;36:i39–52.

78. Fooks GJ, Godziewski C. The World Health Organization, Corporate Power, and the Prevention and Management of Conflicts of Interest in Nutrition Policy Comment on “Towards Preventing and Managing Conflict of Interest in Nutrition Policy? An Analysis of Submissions to a Consultati. Int J Health Policy Manag [Internet]. 2022;11:228–32. Available from: https://www.scopus.com/inward/record.uri?eid=2-s2.0-85125551514&doi=10.34172%2Fijhpm.2020.156&partnerID=40&md5=3a84e743cc8ba9c4de5b74a23cea8dbc

79. Kadandale S, Marten R, Smith R, S. K, R. M. The palm oil industry and noncommunicable diseases. Bull World Health Organ [Internet]. 2019;97:118–28. Available from: https://www.who.int/bulletin/volumes/97/2/18-220434.pdf?ua=1

80. Mialon M, Corvalan C, Cediel G, Scagliusi FBBFB, Reyes M. Food industry political practices in Chile: “the economy has always been the main concern.” Global Health [Internet]. 2020;16. Available from: https://www.scopus.com/inward/record.uri?eid=2-s2.0-85094114467&doi=10.1186%2Fs12992-020-00638-4&partnerID=40&md5=5f40dc19b429497220ed4eb238798d2a

81. Mialon M, Ho M, Carriedo A, Ruskin G, Crosbie E. Beyond nutrition and physical activity: food industry shaping of the very principles of scientific integrity. Global Health. 2021;17.

82. Madureira Lima J, Galea S. The Corporate Permeation Index – A tool to study the macrosocial determinants of Non-Communicable Disease. SSM Popul Health. 2019;7.

83. Steele S, Sarcevic L, Ruskin G, Stuckler D. Confronting potential food industry ‘front groups’: case study of the international food information Council’s nutrition communications using the UCSF food industry documents archive. Global Health [Internet]. 2022;18. Available from: https://www.scopus.com/inward/record.uri?eid=2-s2.0-85124620522&doi=10.1186%2Fs12992-022-00806-8&partnerID=40&md5=236e555adb516f8c68e688a1099a19bd

84. Ramsbottom A, van Schalkwyk MCI, Carters-White L, Benylles Y, Petticrew M. Food as harm reduction during a drinking session: reducing the harm or normalising harmful use of alcohol? A qualitative comparative analysis of alcohol industry and non-alcohol industry-funded guidance. Harm Reduct J [Internet]. 2022;19:66. Available from: http://www.harmreductionjournal.com/home/

85. Watts C, Burton S, Freeman B, C. W, S. B, Watts C, et al. ‘The last line of marketing’: Covert tobacco marketing tactics as revealed by former tobacco industry employees. Glob Public Health [Internet]. 2021;16:1000–13. Available from: http://www.tandf.co.uk/journals/titles/17441692.asp

86. Mialon M. An overview of the commercial determinants of health. Global Health [Internet]. 2020 [cited 2021 May 15];16:74. Available from: /pmc/articles/PMC7433173/

87. Backholer K, Baum F, Finlay SM, Friel S, Giles-Corti B, Jones A, et al. Australia in 2030: what is our path to health for all? Medical Journal of Australia [Internet]. 2021;214:S5–40. Available from: https://www.scopus.com/inward/record.uri?eid=2-s2.0-85105030886&doi=10.5694%2Fmja2.51020&partnerID=40&md5=b0e8f243e5557cdab73f9165d726c401

88. Kasture A, Vandevijvere S, Robinson E, Sacks G, Swinburn B, A. K, et al. Benchmarking the commitments related to population nutrition and obesity prevention of major food companies in New Zealand. Int J Public Health [Internet]. 2019;64:1147–57. Available from: http://ovidsp.ovid.com/ovidweb.cgi?T=JS&PAGE=reference&D=emed20&NEWS=N&AN=628462909

89. Sacks G, Robinson E, Cameron AJ, Vanderlee L, Vandevijvere S, Swinburn B. Benchmarking the nutrition-related policies and commitments of major food companies in Australia, 2018. Int J Environ Res Public Health [Internet]. 2020;17:1–23. Available from: https://www.mdpi.com/1660-4601/17/17/6118/pdf

90. Stubbs T. Commercial determinants of youth smoking in ASEAN countries: A narrative review of research investigating the influence of tobacco advertising, promotion, and sponsorship. Tob Induc Dis [Internet]. 2021;19:1–14. Available from: http://www.tobaccoinduceddiseases.org/Commercial-determinants-of-youth-smoking-in-ASEAN-countries-A-narrative-review-of,139124,0,2.html

91. Baker P, Santos T, Neves PA, Machado P, Smith J, Piwoz E, et al. First-food systems transformations and the ultra-processing of infant and young child diets: The determinants, dynamics and consequences of the global rise in commercial milk formula consumption. Matern Child Nutr [Internet]. 2021;17:e13097. Available from: http://onlinelibrary.wiley.com/journal/10.1111/(ISSN)1740-8709

92. Gillespie D, Hatchard J, Squires H, Gilmore A, Brennan A. Conceptualising changes to tobacco and alcohol policy as affecting a single interlinked system. BMC Public Health [Internet]. 2021;21:17. Available from: https://www.scopus.com/inward/record.uri?eid=2-s2.0-85098644699&doi=10.1186%2Fs12889-020-10000-3&partnerID=40&md5=e5046721715e4d0dfd126d66c88329f6

93. Wood B, Williams O, Nagarajan V, Sacks G. Market strategies used by processed food manufacturers to increase and consolidate their power: a systematic review and document analysis. Global Health [Internet]. 2021;17:17. Available from: http://www.globalizationandhealth.com/

94. Peres MA, Macpherson LMDD, Weyant RJ, Daly B, Venturelli R, Mathur MR, et al. Oral diseases: a global public health challenge. The Lancet [Internet]. 2019;394:249–60. Available from: http://www.journals.elsevier.com/the-lancet/

95. Hessari NM, Bertscher A, Critchlow N, Fitzgerald N, Knai C, Stead M, et al. Recruiting the “heavy-using loyalists of tomorrow”: An analysis of the aims, effects and mechanisms of alcohol advertising, based on advertising industry evaluations. Int J Environ Res Public Health [Internet]. 2019;16:1–17. Available from: https://www.scopus.com/inward/record.uri?eid=2-s2.0-85074139312&doi=10.3390%2Fijerph16214092&partnerID=40&md5=71513da90629d98e92e1c0eeae4cbb3f

96. Kickbusch I, Allen L, Franz C. The commercial determinants of health. Lancet Glob Health [Internet]. 2016 [cited 2021 May 9];4:e895–6. Available from: http://www.who.int/dg/speeches/2013/

97. Gerritsen S, Sing F, Lin K, Martino F, Backholer K, Culpin A, et al. The Timing, Nature and Extent of Social Media Marketing by Unhealthy Food and Drinks Brands During the COVID-19 Pandemic in New Zealand. Front Nutr [Internet]. 2021;8. Available from: https://www.scopus.com/inward/record.uri?eid=2-s2.0-85102938343&doi=10.3389%2Ffnut.2021.645349&partnerID=40&md5=65acfb958cad416bfc974c83ba97fb41

98. McCarthy S, Thomas S, Pitt H, Daube M, Cassidy R. ‘It’s a tradition to go down to the pokies on your 18th birthday’ – the normalisation of gambling for young women in Australia. Aust N Z J Public Health [Internet]. 2020;44:376–81. Available from: https://www.scopus.com/inward/record.uri?eid=2-s2.0-85091146395&doi=10.1111%2F1753-6405.13024&partnerID=40&md5=7092d70312d7104b43a715943a9c898b

99. Maani N, Collin J, Friel S, Gilmore AB, McCambridge J, Robertson L, et al. The need for a conceptual understanding of the macro and meso commercial determinants of health inequalities. Eur J Public Health [Internet]. 2021;31:674–5. Available from: http://ovidsp.ovid.com/ovidweb.cgi?T=JS&PAGE=reference&D=emed22&NEWS=N&AN=635351225

100. Petticrew M, Maani N, Pettigrew L, Rutter H, Van Schalkwyk MCC, M. P, et al. Dark Nudges and Sludge in Big Alcohol: Behavioral Economics, Cognitive Biases, and Alcohol Industry Corporate Social Responsibility. Milbank Quarterly [Internet]. 2020;98:1290–328. Available from: http://onlinelibrary.wiley.com/journal/10.1111/(ISSN)1468-0009

101. Madden M, McCambridge J. Alcohol marketing versus public health: David and Goliath? Global Health. 2021;17.

102. Mialon M, Jaramillo Á, Caro P, Flores M, González L, Gutierrez-Gómez Y, et al. Involvement of the food industry in nutrition conferences in Latin America and the Caribbean. Public Health Nutr. 2021;24:1559–65.

103. Maani N, van Schalkwyk MCI, Filippidis FT, Knai C, Petticrew M. Manufacturing doubt: Assessing the effects of independent vs industry-sponsored messaging about the harms of fossil fuels, smoking, alcohol, and sugar sweetened beverages. SSM Popul Health [Internet]. 2022;17. Available from: https://www.scopus.com/inward/record.uri?eid=2-s2.0-85122512838&doi=10.1016%2Fj.ssmph.2021.101009&partnerID=40&md5=6bb81ee048201ee7b571dbfd0906fbae

104. Passini CSM, Cavalcanti MB, Ribas SA, de Carvalho CMP, Bocca C, Lamarca F. Conflict of Interests in the Scientific Production on Vitamin D and COVID-19: A Scoping Review. Front Public Health [Internet]. 2022;10. Available from: https://www.scopus.com/inward/record.uri?eid=2-s2.0-85134982503&doi=10.3389%2Ffpubh.2022.821740&partnerID=40&md5=18bd911940a964f74588d32277577685

105. Buse K, Mialon M, Jones A. Thinking Politically About UN Political Declarations: A Recipe for Healthier Commitments—Free of Commercial Interests. Int J Health Policy Manag [Internet]. 2022;11:1208–11. Available from: https://www.scopus.com/inward/record.uri?eid=2-s2.0-85135208447&doi=10.34172%2Fijhpm.2021.92&partnerID=40&md5=10f69f23f7b53b81a95bc3339f34f37f

106. Mendly-Zambo Z, Raphael D, Taman A. Take the money and run: how food banks became complicit with Walmart Canada’s hunger producing employment practices. Crit Public Health [Internet]. 2021;00:1–12. Available from: 10.1080/09581596.2021.1955828

107. Kenworthy NJ. Crowdfunding and global health disparities: an exploratory conceptual and empirical analysis. Global Health [Internet]. 2019;15:71. Available from: https://www.scopus.com/inward/record.uri?eid=2-s2.0-85075763795&doi=10.1186%2Fs12992-019-0519-1&partnerID=40&md5=741b59de63218030767d7780a71c1242

108. Wood B, McCoy D, Baker P, Williams O, Sacks G. The double burden of maldistribution: a descriptive analysis of corporate wealth and income distribution in four unhealthy commodity industries. Crit Public Health [Internet]. 2023 [cited 2023 Apr 20];33:135–47. Available from: https://www.tandfonline.com/doi/abs/10.1080/09581596.2021.2019681

109. Rochford C, Tenneti N, Moodie R. Reframing the impact of business on health: The interface of corporate, commercial, political and social determinants of health. BMJ Glob Health [Internet]. 2019 [cited 2021 Sep 16];4:e001510. Available from: https://www.scopus.com/inward/record.uri?eid=2-s2.0-85071114198&doi=10.1136%2Fbmjgh-2019-001510&partnerID=40&md5=c068de6a0e6ebfc5a2664a4c3bf45450

110. The Lancet. Unravelling the commercial determinants of health. In: Maani N, Pettigrew M, Galea S, editors. The Lancet. Oxford Univ Press; 2023. p. 1131.

111. Rose N, Reeve B, Charlton K. Barriers and Enablers for Healthy Food Systems and Environments: The Role of Local Governments. Curr Nutr Rep [Internet]. 2022;11:82–93. Available from: http://www.springer.com/new+%26+forthcoming+titles+%28default%29/journal/13668?changeHeader

112. Liber AC. Using Regulatory Stances to See All the Commercial Determinants of Health. Milbank Quarterly [Internet]. 2022; Available from: http://onlinelibrary.wiley.com/journal/10.1111/(ISSN)1468-0009

113. Loewenson R. Rethinking the Paradigm and Practice of Occupational Health in a World Without Decent Work: A Perspective From East and Southern Africa. New Solutions [Internet]. 2021;31:107–12. Available from: http://ovidsp.ovid.com/ovidweb.cgi?T=JS&PAGE=reference&D=emed22&NEWS=N&AN=635102021

114. Montiel I, Park J, Husted BW, Velez-Calle A. Tracing the connections between international business and communicable diseases. J Int Bus Stud [Internet]. 2022; Available from: https://www.scopus.com/inward/record.uri?eid=2-s2.0-85126885873&doi=10.1057%2Fs41267-022-00512-y&partnerID=40&md5=d0e830c2680d8f506200dc887087b27d

115 consumer. Cambridge Dictionary.

116. worker. Cambridge Dictionary.

117. Appendix E to Part 26 - Individual Determinations of Social and Economic Disadvantage [Internet]. U.S. Department of Transporation. [cited 2023 Nov 5]. Available from: https://www.transportation.gov/civil-rights/appendix-e-part-26-individual-determinations-social-and-economic-disadvantage

118. Frank J, Mustard C, Smith P, Siddiqi A, Cheng Y, Burdorf A, et al. Work as a social determinant of health in high-income countries: past, present, and future. The Lancet [Internet]. 2023;402:1357–67. Available from: 10.1016/S0140-6736(23)00871-1

119. Eisenberg-Guyot J, Prins SJ, Muntaner C. Free agents or cogs in the machine? Classed, gendered, and racialized inequities in hazardous working conditions. Am J Ind Med. 2022;65:92–104.

120. Mohai P, Pellow D, Roberts JT. Environmental justice. Annu Rev Environ Resour. 2009;34:405–30.

121. DuPont-Reyes MJ, Hernandez-Munoz JJ, Tang L. TV Advertising, Corporate Power, and Latino Health Disparities. Am J Prev Med [Internet]. 2022;63:496–504. Available from: https://www.scopus.com/inward/record.uri?eid=2-s2.0-85132157505&doi=10.1016%2Fj.amepre.2022.04.017&partnerID=40&md5=fcb2ff4891b741d88f37c7b64fc9c47e

122. N. R, B. R, Charlton K. AO - Rose NO https://orcid.org/0000-0002-2640-0198, Rose N, Reeve B, Charlton K. Barriers and Enablers for Healthy Food Systems and Environments: The Role of Local Governments. Curr Nutr Rep [Internet]. 2022;11:82–93. Available from: https://www.scopus.com/inward/record.uri?eid=2-s2.0-85124667572&doi=10.1007%2Fs13668-022-00393-5&partnerID=40&md5=24fc56940ecb34b55442d31adfd30a81

123. Ireland R, Bunn C, Reith G, Philpott M, Capewell S, Boyland E, et al. Commercial determinants of health: Advertising of alcohol and unhealthy foods during sporting events. Bull World Health Organ [Internet]. 2019;97:290–5. Available from: https://www.who.int/bulletin/volumes/97/4/18-220087.pdf

124. Klein DE, Lima JM. The Prison Industrial Complex as a Commercial Determinant of Health. Am J Public Health. 2021;111:1750–2.

125. Hyder AA, Werbick M, Scannelli L, Paichadze N. The COVID-19 pandemic exposes another commercial determinant of health: The global firearm industry. Glob Health Sci Pract [Internet]. 2021;9:264–7. Available from: www.ghspjournal.org

126. Jamieson L, Kearns C, Ankeny R, Hedges J, Thomson WMM, L. J, et al. Neoliberalism and Indigenous oral health inequalities: a global perspective. Community Dent Health [Internet]. 2021;38:44–7. Available from: http://ovidsp.ovid.com/ovidweb.cgi?T=JS&PAGE=reference&D=emed22&NEWS=N&AN=634141422

127. Diderichsen F, Dahlgren G, Whitehead M. Beyond “commercial determinants”: Shining a light on privatization and political drivers of health inequalities. Eur J Public Health [Internet]. 2021;31:672–3. Available from: http://ovidsp.ovid.com/ovidweb.cgi?T=JS&PAGE=reference&D=emed22&NEWS=N&AN=635351125

128. Hill SE, Johns P, Nakkash RT, Collin J. From silos to policy coherence: tobacco control, unhealthy commodity industries and the commercial determinants of health. Tob Control [Internet]. 2022;31:322–7. Available from: https://www.scopus.com/inward/record.uri?eid=2-s2.0-85125691950&doi=10.1136%2Ftobaccocontrol-2021-057136&partnerID=40&md5=eaf9dd75ae1ce88ae3e426ec832340ba

129. Boatwright M, Lawrence M, Russell C, Russ K, McCoy D, Baker P. The Politics of Regulating Foods for Infants and Young Children: A Case Study on the Framing and Contestation of Codex Standard-Setting Processes on Breast-Milk Substitutes. Int J Health Policy Manag [Internet]. 2022;11:2422–39. Available from: http://ovidsp.ovid.com/ovidweb.cgi?T=JS&PAGE=reference&D=emexa&NEWS=N&AN=636975055

130. McCambridge J, Coleman R, McEachern J. Public health surveillance studies of alcohol industry market and political strategies: A systematic review. J Stud Alcohol Drugs [Internet]. 2019 [cited 2023 Feb 20];80:149–57. Available from: https://www.jsad.com/doi/10.15288/jsad.2019.80.149

131. Legg T, Hatchard J, Gilmore AB. The Science for Profit Model—How and why corporations influence science and the use of science in policy and practice. PLoS One [Internet]. 2021 [cited 2023 Feb 20];16:e0253272. Available from: https://journals.plos.org/plosone/article?id=10.1371/journal.pone.0253272

132. World Benchmarking Alliance. Methodology for the Food and Agriculture Benchmark. 2021;

133. Sacks G, Vanderlee L, Robinson E, Vandevijvere S, Cameron AJ, Ni Mhurchu C, et al. BIA-Obesity (Business Impact Assessment—Obesity and population-level nutrition): A tool and process to assess food company policies and commitments related to obesity prevention and population nutrition at the national level. Obesity Reviews. 2019;20:78–89.

134. Robertson C. The Money Blind: How to Stop Industry Bias in Biomedical Science, Without Violating the First Amendment. American Journal of Law and Medicine 358 [Internet]. 2011; Available from: https://scholarship.law.bu.edu/faculty_scholarship/1117

135. Israel BA, Schulz AJ, Parker EA, Becker AB. Community-based participatory research: Policy recommendations for promoting a partnership approach in health research. Education for Health. 2001;14:182–97.

