## Supplementary Material for "Corporate activities that influence population health: A scoping review and qualitative synthesis to develop the HEALTH-CORP typology"

**Additional File**

### Appendix 1: Search Strategy Used in Review

**Databases:** Scopus, OVID Medline, Ovid Embase, Ovid Global Health

Searched from inception on Sept 13, 2022

| Database | Search String |
| --- | --- |
| Scopus | TITLE-ABS-KEY ((( corporat* OR commercial*) AND determinant* ) OR "corporate political activity" OR "corporate political activities") AND ( health* OR disease* OR wellbeing OR "well being" OR morbidit* OR mortalit* OR "life expectanc* " OR DALY OR DALYs OR "disability-adjusted life year" OR "disability-adjusted life years" OR QALY OR QALYs OR "quality-adjusted life year" OR "quality-adjusted life years") |
| OVID Medline(R) | ((( corporat* OR commercial*) AND determinant*) OR "corporate political activity" OR "corporate political activities") AND ( health* OR disease* OR wellbeing OR "well being" OR morbidit* OR mortalit* OR "life expectanc*" OR DALY OR DALYs OR "disability-adjusted life year" OR "disability-adjusted life years" OR QALY OR QALYs OR "quality-adjusted life year" OR "quality-adjusted life years").mp. [mp=title, abstract, original title, name of substance word, subject heading word, floating sub-heading word, keyword heading word, organism supplementary concept word, protocol supplementary concept word, rare disease supplementary concept word, unique identifier, synonyms]. |
| Embase | (((corporat* OR commercial*) AND determinant*) OR "corporate political activity" OR "corporate political activities") AND ( health* OR disease* OR wellbeing OR "well being" OR morbidit* OR mortalit* OR "life expectanc*" OR DALY OR DALYs OR "disability-adjusted life year" OR "disability-adjusted life years" OR QALY OR QALYs OR "quality-adjusted life year" OR "quality-adjusted life years").mp. [mp=title, abstract, heading word, drug trade name, original title, device manufacturer, drug manufacturer, device trade name, keyword heading word, floating subheading word, candidate term word] |
| Global Health | ((( corporat* OR commercial*) AND determinant*) OR "corporate political activity" OR "corporate political activities") AND ( health* OR disease* OR wellbeing OR "well being" OR morbidit* OR mortalit* OR "life expectanc*" OR DALY OR DALYs OR "disability-adjusted life year" OR "disability-adjusted life years" OR QALY OR QALYs OR "quality-adjusted life year" OR "quality-adjusted life years").mp. [mp=abstract, title, original title, broad terms, heading words, identifiers, cabicodes] |

### Appendix 2: Table Describing the Characteristics of Included Articles

| Author(s) (Year) | Title | Type of Article | Industry/ies | Regions | Research-Specific Funding Reported? (Y/N) | Type of Funder | Reported Conflict of Interest? (Y/N)^a^ | Open-Access?  (Y/N) |
| --- | --- | --- | --- | --- | --- | --- | --- | --- |
| Millar, J. (2013) | The Corporate Determinants of Health: How Big Business Affects Our Health, and the Need for Government Action! | Commentary | General, with focus on food and beverage | Canada | N | - | N | Y |
| Kickbusch, I. Allen, L., Franz, C. (2016) | The commercial determinants of health | Commentary | General | - | N | - | N | Y |
| Buse, K., Tanaka, S., Hawkes, S (2017) | Healthy people and healthy profits? Elaborating a conceptual framework for governing the commercial determinants of non-communicable diseases and identifying options for reducing risk exposure | Conceptual | Tobacco, ultra-processed foods, alcohol | Global | Y | Educational Institution | N | Y |
| Knai, C. et al. (2018) | Systems Thinking as a Framework for Analyzing Commercial Determinants of Health | Conceptual | General | - | Y | Philanthropic | N | Y |
| McKee, M, Stuckler, D. (2018) | Revisiting the corporate and commercial determinants of health | Conceptual | General | - | N | - | N | Y |
| Wiist, W. (2019) | Mechanisms Underlying Corporations as Determinants of Health | Response | General, focus on tobacco | - | N | - | Y | Y |
| Brown, T. (2019) | Legislative capture: A critical consideration in the commercial determinants of public health | Conceptual | General, with focus on alcohol | Australia | N | - | N | N |
| Ireland, R., Chambers, S., Bunn, C. (2019) | Exploring the relationship between Big Food corporations and professional sports clubs: a scoping review | Review | Food and beverage | Global | Y | Educational Institution | N | Y |
| Toebes, B.,  Patterson, D. (2019) | Human rights and Non-Communicable Diseases: Controlling Tobacco and Promoting Healthy Diets (Book Chapter) | Conceptual | General, focus on tobacco and food and beverage | Global | N | - | N | N |
| Hessari, N. et al. (2019) | Recruiting the “heavy-using loyalists of tomorrow”: An analysis of the aims, effects and mechanisms of alcohol advertising, based on advertising industry evaluations | Qualitative | Alcohol | United Kingdom | Y | Educational Institution | N | Y |
| Peres, M et al. (2019) | Oral diseases: a global public health challenge | Conceptual | Food and beverage | Global | N | - | N | Y |
| Battams, S. & Townsend, B. (2019) | Power asymmetries, policy incoherence and noncommunicable disease control - a qualitative study of policy actor views | Qualitative | General, focus on tobacco, alcohol, food and beverage | Global, Switzerland, Australia, Malaysia | Y | Educational Institution | N | N |
| Madureira Lima, J. & Galea, S. (2019) | The Corporate Permeation Index – A tool to study the macrosocial determinants of Non-Communicable Disease | Scale Development | General | Global | N | - | N | Y |
| Hessari, N. et al. (2019) | Public Meets Private: Conversations Between Coca-Cola and the CDC | Qualitative | Beverage | United States | Y | Philanthropic | Y | Y |
| Ireland, R. et al. (2019) | Commercial determinants of health: Advertising of alcohol and unhealthy foods during sporting events | Conceptual | Alcohol, food and beverage | United Kingdom | Y | Government Entity | Y | Y |
| Kadandale, S., Marten, R., Smith, R. (2019) | The palm oil industry and noncommunicable diseases | Conceptual | Food (palm oil) | Global | N | - | N | Y |
| Kasture, A et al. (2019) | Benchmarking the commitments related to population nutrition and obesity prevention of major food companies in New Zealand | Scale Application | Food and beverage | New Zealand | N | - | N | Y |
| Fooks, G. et al. (2019) | Corporations' use and misuse of evidence to influence health policy: A case study of sugar-sweetened beverage taxation | Qualitative | Beverage | South Africa | N | - | N | Y |
| Kenworthy, N. (2019) | Crowdfunding and global health disparities: an exploratory conceptual and empirical analysis | Conceptual | Crowdfunding | Global | Y | Educational Institution | N | Y |
| Rochford, C., Tenneti, N., Moodie, R. (2019) | Reframing the impact of business on health: the interface of corporate, commercial, political and social determinants of health | Commentary | General | - | N | - | N | Y |
| Mialon et al. (2020) | ‘We must have a sufficient level of profitability’: food industry submissions to the French parliamentary inquiry on industrial food | Qualitative | Food & beverage | France | Y | Government entity | N | N |
| Eastmure, E., Cummins, S., Sparks, L. (2020) | Non-market strategy as a framework for exploring commercial involvement in health policy: A primer | Conceptual | General |  | N |  | N | Y |
| Maani, N., Abdalla, S., Galea, S. (2020) | The firearm industry as a commercial determinant of health | Commentary | Firearm | United States | N | - | N | N |
| Lacy-Nicholas, J., Scrinis, G., Carey, R. (2020) | The politics of voluntary self-regulation: Insights from the development and promotion of the Australian Beverages Council's Commitment | Qualitative | Beverage | Australia | N | - | N | Y |
| Brown, T (2020) | Public health vs alcohol industry compliance laws: A case of regulatory capture? | Conceptual | Alcohol | Australia | N | - | N | N |
| Sacks, G et al. (2020) | Benchmarking the nutrition-related policies and commitments of major food companies in Australia, 2018 | Scale Application | Food and beverage | Australia | N | - | Y | Y |
| Hastings et al. (2020) | Selling second best: how infant formula marketing works | Qualitative | Baby food | United Kingdom, Continental Europe, North America, Australia and New Zealand | Y | International Governance Organization | N | Y |
| De Lacy-Vawdon, C. & Livingstone, C. (2020) | Defining the commercial determinants of health: a systematic review | Review | General | - | N | - | N | Y |
| Mialon, M., Crosbie, E., Sacks, G. (2020) | Mapping of food industry strategies to influence public health policy, research and practice in South Africa | Qualitative | Food and beverage | South Africa | Y | Philanthropic | N | Y |
| Mialon, M. et al. (2020) | Arguments used by trade associations during the early development of a new front-of-pack nutrition labelling system in Brazil | Qualitative | Food and beverage | Brazil | Y | Government Entity | N | N |
| Hill, S., Friel, S. (2020) | ‘As Long as It Comes off as a Cigarette Ad, Not a Civil Rights Message’: Gender, Inequality and the Commercial Determinants of Health | Conceptual | Tobacco, alcohol |  | N |  | N | Y |
| Mialon, M. (2020) | An overview of the commercial determinants of health | Review | General | - | N | - | N | Y |
| Tanrikulu, H. et al. (2020) | Corporate political activity of the baby food industry: The example of Nestlé in the United States of America | Qualitative | Baby food | United States | N | - | N | Y |
| Hoe, C. et al. (2020) | Drink, but don't drive? The alcohol industry's involvement in global road safety | Qualitative | Alcohol | Global | Y | International Governance Organization | N | Y |
| Mialon, M. (2020) | Food industry political practices in Chile: “the economy has always been the main concern” | Qualitative | Food and beverage | Chile | Y | Educational Institution; Government Entity | N | Y |
| Lauber et al., (2020) | Non-communicable disease governance in the era of the sustainable development goals: A qualitative analysis of food industry framing in WHO consultations | Qualitative | Food and beverage | Global | N | - | N | Y |
| Jamieson, L., Gibson, B., Thomson, W. (2020) | Oral health inequalities and the corporate determinants of health: A commentary | Commentary | Food and beverage, alcohol, tobacco | - | N | - | N | Y |
| Karim, Kruger & Hofman (2020) | Industry strategies in the parliamentary process of adopting a sugar-sweetened beverage tax in South Africa: a systematic mapping | Qualitative | Beverage | South Africa | Y | Government entity | N | Y |
| Mialon et al. (2020) | “The architecture of the state was transformed in favour of the interests of companies”: corporate political activity of the food industry in Colombia | Qualitative | Food and beverage | Colombia | Y | Educational Institution; Government Entity | N | Y |
| Petticrew et al. (2020) | Dark Nudges and Sludge in Big Alcohol: Behavioral Economics, Cognitive Biases, and Alcohol Industry Corporate Social Responsibility | Qualitative | Alcohol | - | N | - | N | Y |
| McCarthy et al. (2020) | ‘It’s a tradition to go down to the pokies on your 18th birthday’ – the normalisation of gambling for young women in Australia | Qualitative | Gambling | Australia | Y | Government Entity | N | Y |
| Howse, E et al. (2021) | Air pollution and the noncommunicable disease prevention agenda: Opportunities for public health and environmental science | Commentary | General | - | N | - | N | Y |
| van Schalkwyk, M et al. (2021) | A public health approach to gambling regulation: countering powerful influences | Commentary | Gambling | United Kingdom | N | - | Y | Y |
| Cossez, E., Baker, P., Mialon, M. (2021) | ‘The second mother’: How the baby food industry captures science, health professions and civil society in France | Qualitative | Baby food | France | N | - | N | Y |
| Madden, M. & McCambridge, J. (2021) | Alcohol marketing versus public health: David and Goliath? | Commentary | Alcohol | Global | N | - | N | Y |
| Fisher, L et al. (2021) | Barriers and opportunities to restricting marketing of unhealthy foods and beverages to children in Nepal: a policy analysis | Qualitative | Food and beverage | Nepal | Y | Educational Institution; Government Entity | N | Y |
| Stubbs, T. (2021) | Commercial determinants of youth smoking in ASEAN countries: A narrative review of research investigating the influence of tobacco advertising, promotion, and sponsorship | Review | Tobacco | ASEAN (Association of Southeast Asian Nations) | Y | Educational Institution | N | Y |
| Mialon, M et al. (2021) | Beyond nutrition and physical activity: food industry shaping of the very principles of scientific integrity | Qualitative | General, food and beverage | - | Y | Philanthropic | N | Y |
| Wood, B., Baker, P., Sacks, G. (2021) | Conceptualising the Commercial Determinants of Health Using a Power Lens: A Review and Synthesis of Existing Frameworks | Review | General | - | N | - | N | Y |
| Chavez-Ugalde, Y et al. (2021) | Conceptualizing the commercial determinants of dietary behaviors associated with obesity: A systematic review using principles from critical interpretative synthesis | Review | Food and beverage | - | N | - | N | Y |
| Lauber, K et al. (2021) | Corporate political activity in the context of unhealthy food advertising restrictions across Transport for London: A qualitative case study | Qualitative | Food and beverage | United Kingdom | N | - | Y | Y |
| Freudenberg, N et al. (2021) | Defining Priorities for Action and Research on the Commercial Determinants of Health: A Conceptual Review | Review | General | - | Y | Government entity | N | Y |
| Zenone, M et al. (2021) | How does the British Soft Drink Association respond to media research reporting on the health consequences of sugary drinks? | Qualitative | Beverage | United Kingdom | N | - | N | Y |
| Milaon, M et al. (2021) | Involvement of the food industry in nutrition conferences in Latin America and the Caribbean | Qualitative | Food and beverage | Latin America, Caribbean | N | - | Y | Y |
| Hunt, D. (2021) | How food companies use social media to influence policy debates: A framework of Australian ultra-processed food industry Twitter data | Qualitative | Food and beverage | Australia | N | - | N | Y |
| Wood, B et al. (2021) | Market strategies used by processed food manufacturers to increase and consolidate their power: a systematic review and document analysis | Review | Food and beverage | - | N | - | N | Y |
| Wood, B et al. (2021) | The double burden of maldistribution: a descriptive analysis of corporate wealth and income distribution in four unhealthy commodity industries | Quantitative | Food and beverage, Alcohol, Tobacco, Fossil Fuels | Global, USA | N | - | N | N |
| Lacy-Nichols, J. & Marten, R. (2021) | Power and the commercial determinants of health: Ideas for a research agenda | Commentary | General | - | N | - | N | Y |
| Zenone, M. & Kenworthy, N. (2021) | Pre-emption strategies to block taxes on sugar-sweetened beverages: A framing analysis of Facebook advertising in support of Washington state initiative-1634 | Qualitative | Beverage | United States | N | - | N | Y |
| Loewenson, R. (2021) | Rethinking the Paradigm and Practice of Occupational Health in a World Without Decent Work: A Perspective From East and Southern Africa | Commentary | General, Mining | East and Southern Africa | N | - | N | Y |
| Knai, C et al. (2021) | The case for developing a cohesive systems approach to research across unhealthy commodity industries | Workshop | General | - | Y | Philanthropic | N | Y |
| Mendly-Zambo, Z., Raphael, D., Taman, A. (2021) | Take the money and run: how food banks became complicit with Walmart Canada’s hunger producing employment practices | Conceptual | Retail | Canada | N | - | N | N |
| Hyder, A et al. (2021) | The COVID-19 Pandemic Exposes Another Commercial Determinant of Health: The Global Firearm Industry | Commentary | Firearm | - | Y | Educational Institution | N | Y |
| Campbell, N et al. (2021) | The Gift of Data: Industry-Led Food Reformulation and the Obesity Crisis in Europe | Qualitative | Food and beverage | United Kingdom, Portugal, Ireland, Germany, France | N | - | N | N |
| Zenone, M., Kenworthy, N., Barbic, S. (2021) | The Paradoxical Relationship Between Health Promotion and the Social Media Industry | Commentary | Social media | - | N | - | Y | Y |
| Gerritsen, S et al. (2021) | The Timing, Nature and Extent of Social Media Marketing by Unhealthy Food and Drinks Brands During the COVID-19 Pandemic in New Zealand | Qualitative | Food and beverage | New Zealand | Y | Philanthropic | N | Y |
| Klein, D. & Lima, J. (2021) | The Prison Industrial Complex as a Commercial Determinant of Health | Commentary | Prison | United States | N | - | N | Y |
| McHardy, J. (2021) | The WHO FCTC's lessons for addressing the commercial determinants of health | Commentary | General, tobacco | Global | N | - | Y | Y |
| Russ, K et al. (2021) | What You Don’t Know About the Codex Can Hurt You: How Trade Policy Trumps Global Health Governance in Infant and Young Child Nutrition | Mixed methods | Baby food | Global | Y | Philanthropic | Y | Y |
| Baker, P et al. (2021) | Breastfeeding, first-food systems and corporate power: a case study on the market and political practices of the transnational baby food industry and public health resistance in the Philippines | Qualitative | Baby food | Philippines | Y | International Governance Organization | Y | Y |
| Baker, P et al. (2021) | First-food systems transformations and the ultra-processing of infant and young child diets: The determinants, dynamics and consequences of the global rise in commercial milk formula consumption | Review | Baby food | Global | Y | International Governance Organization | N | Y |
| Baker, P et al. (2021) | Globalization, first-foods systems transformations and corporate power: a synthesis of literature and data on the market and political practices of the transnational baby food industry | Review | Baby food | Global | Y | International Governance Organization | Y | Y |
| Jones et al. (2021) | Disrupting the commercial determinants of health; Chapter 5 in “Australia in 2030 – What is our path to health for all?” (Supplement) | Conceptual | General | Australia | Y | Government Entity | N | Y |
| Maani et al. (2021) | The new WHO Foundation – global health deserves better | Commentary | General | Global | N | - | Y | Y |
| Maani et al. (2021) | The need for a conceptual understanding of the macro and meso commercial determinants of health inequalities | Commentary | General, Alcohol, Tobacco | - | N | - | N | Y |
| Brisbois et al. (2021) | Mining, colonial legacies, and neoliberalism: A political ecology of health knowledge | Conceptual | Extractive | Canada | N | - | Y | Y |
| Jia et al. (2021) | #SupportLocal: how online food delivery services leveraged the COVID-19 pandemic to promote food and beverages on Instagram | Mixed Methods | Food and beverages | Australia, New Zealand, United Kingdom, United States, Canada | N | - | N | Y |
| Mialon et al. (2021) | ‘I had never seen so many lobbyists’: food industry political practices during the development of a new nutrition front-of-pack labelling system in Colombia | Qualitative | Food and beverage | Colombia | Y | Educational Institution; Government Entity | N | Y |
| Dall’Alba & Rocha (2021) | Brazil’s response to COVID-19: commercial determinants of health and regional inequities matter | Commentary | General | Brazil | N | - | N | Y |
| Diderichsen et al. (2021) | Beyond ‘commercial determinants’: shining a light on privatization and political drivers of health inequalities | Commentary | General | Sweden, Denmark | N | - | N | Y |
| Yates et al. (2021) | Trust and responsibility in food systems transformation. Engaging with Big Food: marriage or mirage? | Conceptual | Food and beverage | Global | N | - | N | Y |
| Maani et al. (2021) | The Commercial Determinants of Three Contemporary National Crises: How Corporate Practices Intersect With the  COVID-19 Pandemic, Economic Downturn, and Racial Inequity | Commentary | General | United States | N | - | N | Y |
| Van Schalkwyk et al. (2021) | Our Postpandemic World: What Will It Take to Build a Better Future for People and Planet? | Commentary | General | Global | N | - | N | Y |
| Adams, Rychert, & Wilkins (2021) | Policy inﬂuence and the legalized cannabis industry: learnings from other addictive consumption industries | Conceptual | Cannabis | New Zealand | N | - | Y | N |
| Boatwright et al. (2021) | The Politics of Regulating Foods for Infants and Young Children: A Case Study on the Framing and Contestation of Codex Standard-Setting Processes on Breast-Milk Substitutes | Mixed Methods | Baby food | Global | Y | International Governance Organization | N | Y |
| Watts, Burton & Freman (2021) | ‘The last line of marketing’: Covert tobacco marketing tactics as revealed by former tobacco industry employees | Qualitative | Tobacco | Australia | Y | Philanthropic | N | Y |
| Gillespie et al. (2021) | Conceptualising changes to tobacco and alcohol policy as affecting a single interlinked system | Workshop | Tobacco, alcohol | United Kingdom | Y | Philanthropic; Government Entity; Educational Institution | N | Y |
| Jamieson et al. (2021) | Neoliberalism and Indigenous oral health inequalities: a global perspective | Commentary | Tobacco, food and beverage | Global | N | - | N | N |
| Hill et al. (2021) | From silos to policy coherence: tobacco control, unhealthy commodity industries and the commercial determinants of health | Commentary | Tobacco, General | - | N | - | Y | Y |
| McCarthy, S et al. (2022) | Electronic gambling machine harm in older women: a public health determinants perspective | Qualitative | Gambling | Australia | N | - | Y | N |
| Lee, K et al. (2022) | Measuring the Commercial Determinants of Health and Disease: A Proposed Framework | Scale Development | General | - | Y | Government Entity | N | Y |
| Allen, Wigley, and Homer (2022) | Assessing the association between Corporate Financial Influence and implementation of policies to tackle commercial determinants of non-communicable diseases: A cross-sectional analysis of 172 countries | Quantitative | General | Global | N | - | N | Y |
| Buse, Mialon, Jones (2022) | Thinking politically about UN political declarations: A recipe for healthier commitments – free of commercial interests | Conceptual | General | - | N | - | N | Y |
| Clare, Maani, and Milner (2022) | Meat, money and messaging: How the environmental and health harms of red and processed meat consumption are framed by the meat industry | Qualitative | Food (meat) | United Kingdom | N | - | N | N |
| Barlow & Allen (2022) | The impact of trade and investment agreements on the implementation noncommunicable disease policies, 2014-2019: protocol for a statistical study (preprint) | Quantitative Protocol | Tobacco, alcohol, food and beverage | Global | N | - | N | Y |
| DuPont-Reyes, Hernandez-Munoz, and Tang (2022) | TV advertising, corporate power, and Latino health disparities | Mixed Methods | Alcohol, tobacco, food and beverage, pharmaceutical | United States | N | - | N | N |
| Gokani et al. (2022) | UK Nutrition Research Partnership ‘Hot Topic’ workshop report: A ‘game changer’ for dietary health – addressing the implications of sport sponsorship by food businesses through an innovative interdisciplinary collaboration | Workshop | Food and beverage | United Kingdom | Y | Government Entity | N | Y |
| Hird et al. (2022) | Understanding the long-term policy influence strategies for the tobacco industry: two contemporary case studies | Conceptual | Tobacco | - | Y | Philanthropic | N | Y |
| Montiel et al. (2022) | Tracing the connections between international business and communicable diseases | Conceptual | General | Global | Y | Educational Institution | N | Y |
| Fooks & Godziewski (2022) | The World Health Organization, Corporate Power, and the Prevention and Management of Conflicts of Interest in Nutrition Policy: Comment on “Towards Preventing and Managing Conflict of Interest in Nutrition Policy? An Analysis of Submissions to a Consultation on a Draft WHO Tool” | Commentary | Food and beverage | Global | N | - | N | Y |
| Zenone, Kenworthy, & Maani (2022) | The Social Media Industry as a Commercial Determinant of Health | Commentary | Social media |  | N | - | N | Y |
| Mialon et al. (2022) | Conflicts of interest for members of the U.S. 2020 Dietary Guidelines Advisory Committee | Quantitative | Food and beverage | United States | Y | Philanthropic | N | Y |
| Passini et al. (2022) | Conflict of interests in the scientific production of Vitamin D and COVID-19: A Scoping Review | Review | Food, diagnostics, pharmaceutical | - | Y | Educational Institution | N | Y |
| Ramsbottom et al. (2022) | Food as harm reduction during a drinking session: reducing the harm or normalising harmful use of alcohol? A qualitative comparative analysis of alcohol industry and non-alcohol industry-funded guidance | Qualitative | Alcohol | - | N | - | N | Y |
| Rose, Reeve & Charlton (2022) | Barriers and Enablers for Healthy Food Systems and Environments: The Role of Local Governments | Review | Food and beverage | - | N | - | N | Y |
| Steele et al. (2022) | Confronting potential food industry ‘front groups’: case study of the international food information Council’s nutrition communications using the UCSF food industry documents archive | Qualitative | Food and beverage | Global | Y | Philanthropic | Y | Y |
| De Lacy-Vawdon, Vandenberg, & Livingstone (2022) | Recognising the elephant in the room: the commercial determinants of health | Commentary | General | - | N | - | N | Y |
| Wiist, W. (2022) | The Foundations of Corporate Strategies: Comment on “‘Part of the Solution’: Food Corporation Strategies for Regulatory Capture and Legitimacy” | Commentary | General | United States | N | - | Y | Y |
| Maani et al. (2022) | Manufacturing doubt: Assessing the effects of independent vs industry-sponsored messaging about the harms of fossil fuels, smoking, alcohol, and sugar sweetened beverages | Quantitative | Alcohol, tobacco, fossil fuels, sugar-sweetened beverages | United Kingdom | Y | Government Entity | N | Y |
| Hoe et al. (2022) | Strategies to expand corporate autonomy by the tobacco, alcohol and sugar-sweetened beverage industry: a scoping review of reviews | Review | Tobacco, alcohol, sugar-sweetened beverages | - | Y | Philanthropic | N | Y |
| Freudenberg (2022) | Responding to Food Industry Initiatives to Be “Part of the Solution”  Comment on “‘Part of the Solution’: Food Corporation Strategies for Regulatory Capture and Legitimacy” | Response | Food and beverage | - | N | - | N | Y |
| Kroker-Lobos et al. (2022) | Two countries, similar practices: The political practices of the food industry influencing the adoption of key public health nutrition policies in Guatemala and Panama | Qualitative | Food | Guatemala, Panama | Y | Educational Institution; Government Entity | N | Y |
| Leimbigler et al. (2022) | Social, political, commercial, and corporate determinants of rural health equity in Canada: an integrated framework | Commentary | General | Canada | Y | Educational Institution | N | Y |
| Liber (2022) | Using Regulatory Stances to See All the Commercial Determinants of Health | Conceptual | General, Pharma/Diagnostics, Food & beverage, Housing, Tobacco (E-Cigarettes) | - | N | - | Y | Y |
| Wakefield, Glantz & Appollonio (2022) | Content Analysis of the Corporate Social Responsibility Practices of 9 Major Cannabis Companies in Canada and the US | Qualitative | Cannabis | United States, Canada | Y | Government Entity | N | Y |
| Van Schalkwyk, Hawkins & Pettigrew (2022) | The politics and fantasy of the gambling education discourse: An analysis of gambling industry-funded youth education programmes in the United Kingdom | Qualitative | Gambling | United Kingdom | Y | Government Entity | N | Y |

^a^Articles labelled with ‘N’ in this column either specifically declared that there was no related funding, reported funding that was not specific to the research (e.g., general support for investigators) or information on funding was not available (i.e., not reported). ‘Y’ indicated funding specific to the article in question was reported.

^b^Articles labelled with ‘N’ in this column either specifically declared no competing interests or information on competing interests was not available (i.e., not declared). ‘Y’ indicates that a competing interest was declared.

### Appendix 3: The Corporate Influences on Health (HEALTH-CORP) Typology

| Domains of Corporate Influence | Definition of Domain | Corporate Activities with Potential to Influence Population Health and/or Health Equity^a,b^ | Expected Direction of Health Impact^c^ |
| --- | --- | --- | --- |
| DISTAL DOMAINS | | | |
| Political Practices  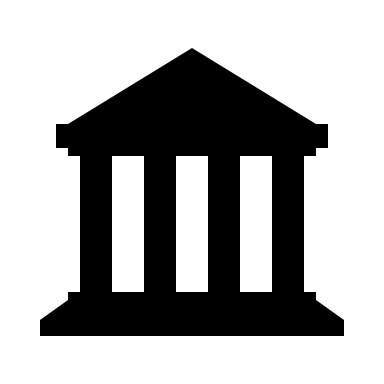 | This domain consists of activities undertaken to influence government policy or processes in ways that are favourable to the commercial entity [1]. | ***Activities related to securing a favourable policy environment:*** | |
|  |  | Engage in political financing | Depends |
|  |  | Engage in bribery (e.g., provide gifts, other incentives to policy makers) | - |
|  |  | Advocate for policies that limit corporate liability for health harms | - |
|  |  | Engage in efforts to develop relationships between corporations and public health institutions (e.g., through public-private partnerships, funding, ‘wine-ing and dining’) | Depends |
|  |  | Advocate for the placement of corporate representatives on regulatory boards and research associations | - |
|  |  | Exploit the use of ‘revolving doors’ (i.e., employees who move between positions in industry and government [2]) | - |
|  |  | ***Activities related to stopping, delaying, or weakening proposed health and related policies:*** | |
|  |  | Take or threaten legal action in response to proposed health policies | - |
|  |  | Advocate for engagement in regulation (e.g., self-regulation, co-regulation) | - |
|  |  | Engage in political lobbying, including through the use of front groups | Depends |
|  |  | Engage in strategies (e.g., introducing ballot measures [3]) to leverage pre-emption (i.e., higher levels of government restrict the jurisdiction of lower levels [4]) | Depends |
|  |  | Leverage trade treaties to challenge health policies | - |
|  |  | Develop industry alliances to oppose proposed policies with a united front | - |
|  |  | Use argumentative strategies to oppose proposed health policies within policy submissions  (e.g., suggest policy is not within the mandate of the regulating institution [5]) | - |
|  |  | Misrepresent evidence or demand unrealistic standards of public health evidence within policy submissions | - |
|  |  | Exploit existing divisions in the public health community | - |
|  |  | Engage in intimidation tactics, including efforts to discredit opposing scientists and policy makers | - |
|  |  | Shift or threaten to shift operations to countries with weaker regulations | - |
|  |  | ***Other political activities:*** |  |
|  |  | Advocate for privatization of public services | Depends |
| Preference & Perception Shaping Practices  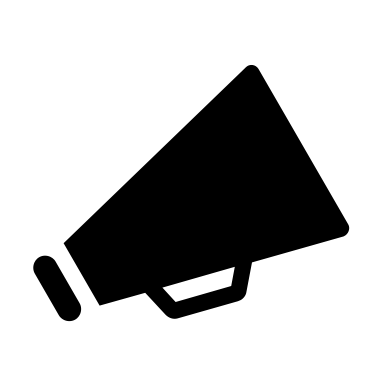 | This domain consists of activities that shape preferences for products and/or influence perceptions about products and their health-related harms [1]. | ***Activities related to promoting products:*** | |
|  |  | Engage in marketing | Depends |
|  |  | Engage in marketing of harmful products that is disproportionately targeted towards disadvantaged groups | - |
|  |  | Leverage pandemics (e.g., COVID-19 [6]) or other disasters to promote products | - |
|  |  | Sponsor sports teams | Depends |
|  |  | ***Activities related to shaping the public debate about products & their health implications:*** | |
|  |  | Engage in health education efforts directed at the public (e.g., alcohol industry’s messaging about the health harms of alcohol consumption during pregnancy [7]) | Depends |
|  |  | Engage in efforts to influence the public’s perception of health policies (e.g., spread misinformation about policies [3]) | - |
|  |  | Reframe the causes of health issues (e.g., suggest that physical activity is more important than diet in weight management [8]), including through the use of front groups | - |
|  |  | Advance the idea of individual responsibility for health | Depends |
|  |  | Acquire, fund, or establish relationships with media companies | - |
|  |  | Employ medical experts or scientists to advance industry interests (e.g., by giving lectures supporting harmful products [9]) | - |
|  |  | ***Activities related to shaping the professional debate about products & their health implications:*** | |
|  |  | Provide funding to professional associations | Depends |
|  |  | Contribute to the development of clinical standards (e.g., cows-milk protein allergy [10]) | Depends |
|  |  | Engage in health education efforts targeted at health care professionals (or health care professional students) | Depends |
|  |  | ***Activities related to the production of evidence and the academic debate about products and their health implications:*** | |
|  |  | Provide funding for research, via provision of funding to universities, scientific conferences, academic journals, scientific awards, think tanks, or through development of industry research institutes | Depends |
|  |  | Suppress or amplify research (e.g., via social media bots [11]) depending on its desirability to industry | - |
|  |  | Obscure conflicts of interest (i.e., relationships between authors and industry [12]) in research | - |
|  |  | Contribute to the development of scientific standards (e.g., principles of scientific integrity [13]) | Depends |
|  |  | Falsify data | - |
| Corporate Social Responsibility Practices 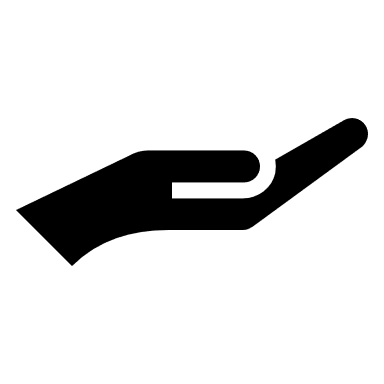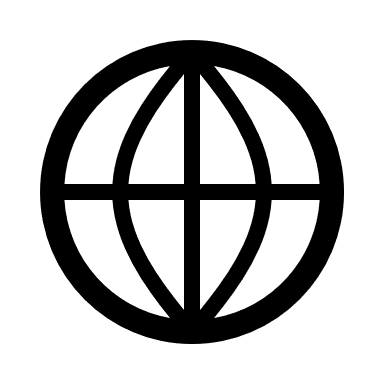 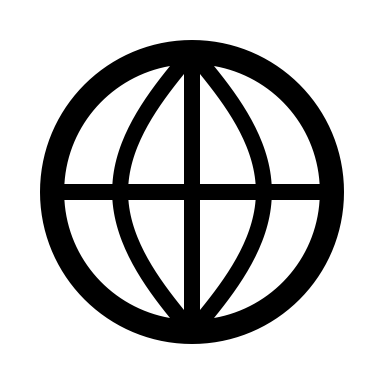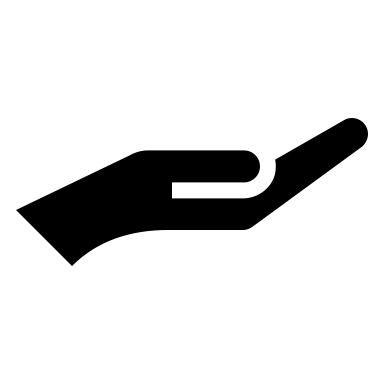 | This domain consists activities undertaken with the stated intention to contribute to society and/or offset environmental, social, or health impacts of previous activities [14]. | Develop or contribute to health promotion programs (e.g., programs to reduce drunk driving [15]), or health charities (e.g., HIV prevention initiatives [9]), including health education efforts | Depends |
|  |  | Engage in other social responsibility initiatives that are relevant to health (e.g., diversity, equity, and inclusion efforts [16]) | Depends |
|  |  | Engage with existing social movements (e.g., the women’s rights movement [17]) | Depends |
|  |  | Reformulate products for health-related reasons (e.g., develop drinks with artificial sweeteners to reduce sugar content [18]) | Depends |
| Economic Practices  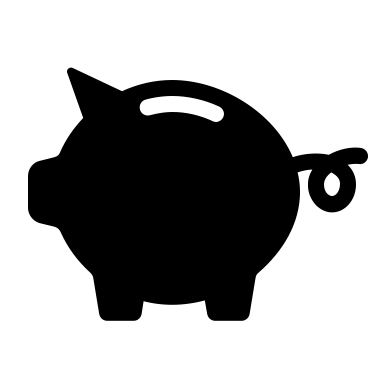 | This domain consists of activities that influence the economy and the distribution of wealth within society [19]. | Engage in fair vs unfair tax practices (e.g., tax evasion, tax avoidance [20]) | +/- |
|  |  | Engage in price fixing of necessary commodities (e.g., food [21]) | - |
|  |  | Contribute to economic growth and related benefits (e.g., improvements in infrastructure, education, healthcare) | + |
|  |  | Contribute to inequitable distributions of wealth and power through ownership and renumeration structures that prioritize the accrual of wealth to certain individuals (i.e., executives, shareholders) over others (i.e., workers) | - |
| PROXIMAL DOMAINS | | | |
| Products & Services  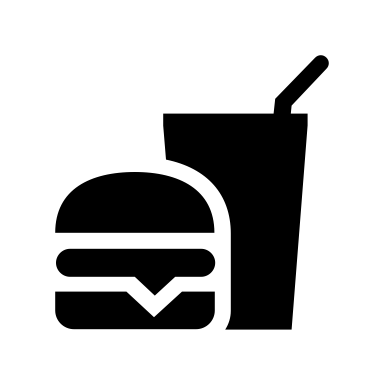 | This domain consists of activities related to the production and sale of products and services [22]. | ***Activities related to the characteristics of products:*** | |
|  |  | Develop, produce, or sell products with harmful (e.g., cigarettes) or salutogenic properties (e.g., vaccines) | +/- |
|  |  | Determine the addictive properties of products | +/- |
|  |  | ***Activities related to the accessibility of products:*** | |
|  |  | Determine the price of products (for e.g., low prices of ultra-processed foods, use of price promotions [23]) | Depends |
|  |  | Determine the proximity and availability of access to products (e.g., hours of operation of gambling outlets [24]) | Depends |
| Employment Practices  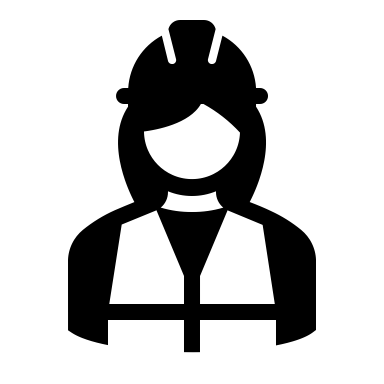 | This domain consists of activities related to the conditions under which employment is provided [25]. | ***Activities related to the characteristics of employment:*** | |
|  |  | Number of employment opportunities | +/- |
|  |  | Determine the adequacy of pay in relation to living requirements | +/- |
|  |  | Determine the stability of employment terms | +/- |
|  |  | ***Activities related to the benefits received through employment:*** | |
|  |  | Determine the provision and quality of medical benefits | +/- |
|  |  | Determine the provision and quality of pension plans | +/- |
|  |  | Determine the provision, length, and paid or unpaid status of parental leave | +/- |
|  |  | Determine the provision and quality of employee wellness programs | +/- |
|  |  | ***Activities related to the conditions of employment:*** |  |
|  |  | Determine the quality of working conditions (e.g., safety of working environments) | +/- |
|  |  | Engage in anti-union activities that prevent or discourage workers from unionizing or engaging in collective bargaining | - |
|  |  | Determine the provision of support for breastfeeding in the workplace | +/- |
|  |  | Determine the provision of opportunities to work remotely and the characteristics of remote work (e.g., organizational support [26]) | +/- |
|  |  | ***Other employment-related activities:*** | |
|  |  | Determine the presence of child labor or forced labor directly or in the supply chain | +/- |
| Environmental Practices  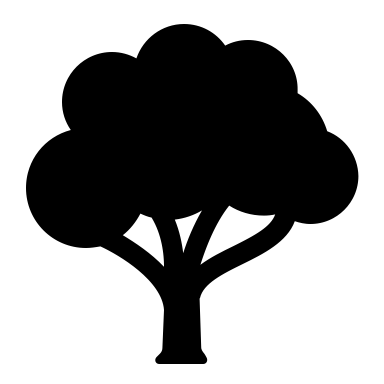 | This domain consists of corporate activities that can influence physical and/or mental health through the impact of the activity on the natural environment [27]. | Use harmful chemicals and pesticides | - |
|  |  | Pollute air | - |
|  |  | Pollute water | - |
|  |  | Produce waste | - |
|  |  | Extract resources (e.g., water [28]) | - |
|  |  | Contribute to deforestation | - |
|  |  | Consume energy | - |
|  |  | Expropriate land for industry activities (e.g., mining [26]) | - |
|  |  | Contribute to greenhouse gas emissions | - |
|  |  | Contribute to biodiversity loss | - |

^a^The corporate activities are described using verbs (e.g., contribute, engage) to draw attention to the active role that corporations play in shaping population health. However, we recognize that there are other actors (e.g., the government) involved that play a role in how these activities are enacted (e.g., federal regulations for parental leave policies).

^b^The expected associated health impact may be direct (such as producing health-harming products) or indirect (such as using argumentative strategies to oppose proposed health policies within policy submissions).

^c^A “+” indicates that the respective activity is expected to have a positive impact on human health. A “-” indicates that the respective activity is expected to have a negative impact on health. A “+/-” indicates that the activity can produce a positive or a negative impact on human health depending on the extent to which the activity is present or absent or its quality. For example, providing medical benefits to employees would likely result in a positive health impact whereas not doing so would likely result in a negative health impact [29]. A “Depends” indicates that whether the impact on human health is positive or negative depends on the specifics of the respective activity. For example, providing funding for a scientific conference may have an indirect positive or negative impact on human health *depending* on whether the funding allows the corporation to influence the agenda of the respective conference.

### Appendix 4: Activities Recorded in One Industry

The following activities were recorded with respect to one industry only and therefore were not included in the HEALTH-CORP typology:

| Corporate Activities with Potential to Influence Population Health and/or Health Equity | Direction of Expected Health Impact^a^ | Industry in Which the Activity Was Discussed |
| --- | --- | --- |
| Determine the presence of socially-isolated employee communities (e.g., migrant or ‘fly-in’ workers) | +/- | Extractive |
| Determine the provision and quality of harm prevention supports | +/- | Gambling |
| Place restrictions on corporate data (making it difficult to assess health harms) | - | Social Media |
| Undermine social safety nets | - | Crowdfunding |
| Engage in unapproved research | - | Food & Beverage |

^a^A “+” indicates that the respective activity is expected to have a positive impact on human health. A “-” indicates that the respective activity is expected to have a negative impact on health. A “+/-” indicates that the activity can produce a positive or a negative impact on human health depending on the extent to which the activity is present or absent or its quality. For example, providing medical benefits to employees would likely result in a positive health impact whereas not doing so would likely result in a negative health impact [29]. A “Depends” indicates that whether the impact on human health is positive or negative depends on the specifics of the respective activity. For example, providing funding for a scientific conference may have an indirect positive or negative impact on human health *depending* on whether the funding allows the corporation to influence the agenda of the respective conference.

### Appendix 5: Industries in Which the Domains of Influence Were Discussed

| Domains of Corporate Influence | General | Food & Beverage | Tobacco | Alcohol | Baby Food | Gambling | Extractive | Pharma and Diagnostics | Firearms | Social Media | Cannabis | Prisons | Retail | Crowd-funding | E-Cigarettes | Housing | Fossil Fuels | Total number of industries identified per domain |
| --- | --- | --- | --- | --- | --- | --- | --- | --- | --- | --- | --- | --- | --- | --- | --- | --- | --- | --- |
| Political Practices |  |  |  |  |  |  |  |  |  |  |  |  |  |  |  |  |  | **9** |
| Preference & Perception Shaping Practices |  |  |  |  |  |  |  |  |  |  |  |  |  |  |  |  |  | **11** |
| Corporate Social Responsibility (CSR) Practices |  |  |  |  |  |  |  |  |  |  |  |  |  |  |  |  |  | **10** |
| Economic Practices |  |  |  |  |  |  |  |  |  |  |  |  |  |  |  |  |  | **7** |
| Products & Services |  |  |  |  |  |  |  |  |  |  |  |  |  |  |  |  |  | **12** |
| Employment Practices |  |  |  |  |  |  |  |  |  |  |  |  |  |  |  |  |  | **5** |
| Environmental Practices |  |  |  |  |  |  |  |  |  |  |  |  |  |  |  |  |  | **3** |
| Total number of domains identified for respective industry | **7** | **7** | **5** | **4** | **6** | **4** | **5** | **3** | **2** | **2** | **2** | **1** | **4** | **1** |  | **1** | **2** | **-** |

**References in Additional File**

1.	Madureira Lima J, Galea S. Corporate practices and health: A framework and mechanisms. Globalization and Health. BioMed Central Ltd.; 2018. pp. 1–12. doi:10.1186/s12992-018-0336-y

2.	Definition of “the revolving door.” In: Collins Dictionary [Internet]. [cited 14 Nov 2023]. Available: https://www.collinsdictionary.com/us/dictionary/english/the-revolving-door

3.	Zenone M, Kenworthy N, M. Z, Zenone M, Kenworthy N. Pre-emption strategies to block taxes on sugar-sweetened beverages: A framing analysis of Facebook advertising in support of Washington state initiative-1634. Glob Public Health. 2021;17: 1854–1867. doi:https://dx.doi.org/10.1080/17441692.2021.1977971

4.	Crosbie E, Schillinger D, Schmidt LA. State Preemption to Prevent Local Taxation of Sugar-Sweetened Beverages. JAMA Intern Med. 2019;179: 291–292. doi:10.1001/JAMAINTERNMED.2018.7770

5.	K. L, R. R, M. M, A. C, Gilmore A.B.  AO  - Lauber KO https://orcid. org/0000-0003-0073-3004, Lauber K, et al. Non-communicable disease governance in the era of the sustainable development goals: A qualitative analysis of food industry framing in WHO consultations. Global Health. 2020;16: 76. doi:https://dx.doi.org/10.1186/s12992-020-00611-1

6.	Gerritsen S, Sing F, Lin K, Martino F, Backholer K, Culpin A, et al. The Timing, Nature and Extent of Social Media Marketing by Unhealthy Food and Drinks Brands During the COVID-19 Pandemic in New Zealand. Front Nutr. 2021;8. doi:10.3389/fnut.2021.645349

7.	Maani N, van Schalkwyk MCI, Filippidis FT, Knai C, Petticrew M. Manufacturing doubt: Assessing the effects of independent vs industry-sponsored messaging about the harms of fossil fuels, smoking, alcohol, and sugar sweetened beverages. SSM Popul Health. 2022;17. doi:10.1016/j.ssmph.2021.101009

8.	Buse K, Tanaka S, Hawkes S. Healthy people and healthy profits? Elaborating a conceptual framework for governing the commercial determinants of non-communicable diseases and identifying options for reducing risk exposure. Globalization and Health. BioMed Central Ltd.; 2017. doi:10.1186/s12992-017-0255-3

9.	Adams PJJ, Rychert M, Wilkins C. Policy influence and the legalized cannabis industry: learnings from other addictive consumption industries. Addiction. 2021;116: 2939–2946. doi:10.1111/add.15483

10.	Baker P, Santos T, Neves PA, Machado P, Smith J, Piwoz E, et al. First-food systems transformations and the ultra-processing of infant and young child diets: The determinants, dynamics and consequences of the global rise in commercial milk formula consumption. Matern Child Nutr. 2021;17: e13097. doi:10.1111/mcn.13097

11.	Steele S, Sarcevic L, Ruskin G, Stuckler D. Confronting potential food industry ‘front groups’: case study of the international food information Council’s nutrition communications using the UCSF food industry documents archive. Global Health. 2022;18. doi:10.1186/s12992-022-00806-8

12.	Thompson DF. Understanding Financial Conflicts of Interest. New England Journal of Medicine. 1993; 573–576. doi:10.1056/NEJM199308193290812

13.	Mialon M, Ho M, Carriedo A, Ruskin G, Crosbie E. Beyond nutrition and physical activity: food industry shaping of the very principles of scientific integrity. Global Health. 2021;17. doi:10.1186/s12992-021-00689-1

14.	Wakefield T, Glantz SA, Apollonio DE, T. W, S.A. G. Content Analysis of the Corporate Social Responsibility Practices of 9 Major Cannabis Companies in Canada and the US. JAMA Netw Open. 2022; E2228088. doi:10.1001/jamanetworkopen.2022.28088

15.	Hoe C, Taber N, Champagne S, Bachani AM. Drink, but don’t drive? The alcohol industry’s involvement in global road safety. Health Policy Plan. 2020;35: 1328–1338. doi:10.1093/heapol/czaa097

16.	Wakefield T, Glantz SA, Apollonio DE, T. W, S.A. G. Content Analysis of the Corporate Social Responsibility Practices of 9 Major Cannabis Companies in Canada and the US. JAMA Netw Open. 2022; E2228088. doi:10.1001/jamanetworkopen.2022.28088

17.	Hill SE, Friel S. ‘As long as it comes off as a cigarette ad, not a civil rights message’: Gender, inequality and the commercial determinants of health. Int J Environ Res Public Health. 2020;17: 1–19. doi:10.3390/ijerph17217902

18.	Mialon M, Corvalan C, Cediel G, Scagliusi FBBFB, Reyes M. Food industry political practices in Chile: “the economy has always been the main concern.” Global Health. 2020;16. doi:10.1186/s12992-020-00638-4

19.	Baum FE, Sanders DM, Fisher M, Anaf J, Freudenberg N, Friel S, et al. Assessing the health impact of transnational corporations: Its importance and a framework. Global Health. 2016;12: 1–7. doi:10.1186/s12992-016-0164-x

20.	Wood B, McCoy D, Baker P, Williams O, Sacks G. The double burden of maldistribution: a descriptive analysis of corporate wealth and income distribution in four unhealthy commodity industries. Crit Public Health. 2023;33: 135–147. doi:10.1080/09581596.2021.2019681

21.	Mendly-Zambo Z, Raphael D, Taman A. Take the money and run: how food banks became complicit with Walmart Canada’s hunger producing employment practices. Crit Public Health. 2021;00: 1–12. doi:10.1080/09581596.2021.1955828

22.	Knai C, Petticrew M, Capewell S, Cassidy R, Collin J, Cummins S, et al. The case for developing a cohesive systems approach to research across unhealthy commodity industries. BMJ Glob Health. 2021;6. doi:10.1136/bmjgh-2020-003543

23.	Chavez-Ugalde Y, Jago R, Toumpakari Z, Egan M, Cummins S, White M, et al. Conceptualizing the commercial determinants of dietary behaviors associated with obesity: A systematic review using principles from critical interpretative synthesis. Obes Sci Pract. 2021;7: 473–486. doi:10.1002/osp4.507

24.	McCarthy S, Thomas S, Pitt H, Daube M, Cassidy R. ‘It’s a tradition to go down to the pokies on your 18th birthday’ – the normalisation of gambling for young women in Australia. Aust N Z J Public Health. 2020;44: 376–381. doi:10.1111/1753-6405.13024

25.	Occupational Health. In: International Labour Organization [Internet]. [cited 28 Feb 2023]. Available: https://www.ilo.org/safework/areasofwork/occupational-health/lang--en/index.htm

26.	Loewenson R. Rethinking the Paradigm and Practice of Occupational Health in a World Without Decent Work: A Perspective From East and Southern Africa. New Solutions. 2021;31: 107–112. doi:10.1177/10482911211017106

27.	Sattler B. Environmental Health. Policy Polit Nurs Pract. 2003;4: 4–5. doi:10.1177/1527154402239448

28.	Montiel I, Park J, Husted BW, Velez-Calle A. Tracing the connections between international business and communicable diseases. J Int Bus Stud. 2022. doi:10.1057/s41267-022-00512-y

29.	Van Niel MS, Bhatia R, Riano NS, De Faria L, Catapano-Friedman L, Ravven S, et al. The Impact of Paid Maternity Leave on the Mental and Physical Health of Mothers and Children: A Review of the Literature and Policy Implications. Harv Rev Psychiatry. 2020;28: 113–126. doi:10.1097/HRP.0000000000000246
